# Genetic Epidemiological Evidence Indicates the Role of Lipid Control in Omega - 3 – Mediated Cardiovascular Protection

**DOI:** 10.1101/2025.09.03.25333717

**Authors:** Jinhan Yu, Chengjia Li, Zipeng Liu, Yidan Chen, Yao Lei, Dingyang Li, Wen Ma, Yao Wang, Yi Yang, Renzhi Wang, Yong-Fei Wang

**Author notes:** Correspondence to: Yong-Fei Wang. These authors contributed equally (Jinhan Yu, Chengjia Li).

## Abstract

**Background:** The association between omega-3 fatty acids and cardiovascular disease has been examined in populations with established disease. However, the definitive role of omega-3 fatty acids in cardiovascular risk remains inconclusive, and evidence from general populations is still limited. This study aims to examine the causal relationship between omega-3 and cardiovascular disease using large-scale genetic data.

**Methods:** We employed five well-established mendelian randomization (MR) methods to investigate the causal association between omega-3 and cardiovascular events. Two large independent omega-3 GWAS (>110,000 participants each) from general populations were analyzed across 14 cardiovascular and metabolic phenotypes (>50,000 participants each). To understand the direct effect of omega-3 on coronary artery disease (CAD), multivariable MR (MVMR) model and mediation analysis were conducted. Summary-data-based MR (SMR) was applied to identify circulating proteins associated with omega-3 levels.

**Results:** Genetically elevated omega-3 levels were found associated with increased cardiovascular risk across all MR methods, particularly for CAD (*P*_median_ = 2.65E-04), myocardial infarction (*P*_median_ = 1.03E-07) and heart failure (*P*_median_ = 4.84E-03). Higher genetically predicted omega-3 were linked to elevated LDL-C (*P*_median_ = 2.50E-33) and its key component apolipoprotein B (*P*_median_ = 5.60E-08), while reducing triglyceride levels (*P*_median_ = 4.16E-03). Notably, after adjusting for the genetic effect of LDL-C, omega-3 demonstrated a protective effect on CAD (β_combined_ = −0.025, *P*_combined_ = 0.022), which was strengthened by adjusting for both LDL-C and triglycerides. Mediation analysis suggests that omega-3 is likely to increase CAD risk through elevating LDL-C (*P* _median_ = 2.40E-12), highlighting the need for effective LDL-C management to maximize the cardiovascular benefits of omega-3. Circulating proteins, such as ANGPTL3 (*P*_smr_ = 1.22E-27) and PCSK9 (*P*_smr_ = 1.15E-21), were also found positively associated with omega-3 levels. A reanalysis of clinical trial data confirmed that the cardiovascular benefits of omega-3 were closely related to the intensity of LDL-C lowering therapy.

**Conclusions:** Our results suggest that the cardiovascular benefits of omega-3 fatty acids may depend on effective lipid management, particularly LDL-C control. Combining omega-3 supplementation with advanced lipid-lowering therapy may be critical for maximizing their protective impact on cardiovascular health.

## Introduction

Omega-3 fatty acids, a type of polyunsaturated fatty acids (PUFAs), have long been investigated for their potential protective effects on cardiovascular health^1,2^. However, evidence from two large-scale randomized clinical trials (RCTs) remains inconclusive. The Reduction of Cardiovascular Events with Icosapent Ethyl–Intervention Trial (REDUCE-IT, n = 8,179; NCT01492361) reported that high-dose icosapent ethyl (a purified and stable EPA ethyl ester, 4g/d) significantly reduced major cardiovascular events by 25% in patients with hypertriglyceridemia^3^. But concerns have been raised regarding the use of mineral oil as placebo. In contrast, the Long-Term Outcomes Study to Assess Statin Residual Risk with Epanova in High Cardiovascular Risk Patients with Hypertriglyceridemia (STRENGTH, n = 13,078; NCT02104817) trial found no significant cardiovascular benefit with omega-3 CA (a carboxylic acid formulation of EPA and DHA, 4g/d) compared to corn oil over a median follow-up of 42.0 months^4^.

Previous studies were primarily conducted in populations with cardiovascular-related conditions^3,4^, and the conflicting results demonstrated the challenges in investigating a clear causal relationship between omega-3 fatty acids and cardiovascular outcomes. These inconsistencies may result from placebo selection, drug use, dosing regimens, study populations, and the difficulty of controlling numerous confounding factors. Mendelian randomization (MR) offers a way to address these limitations by using genetic variants as instrumental variables (IVs) for exposures. This method reduces bias from potential confounders, as genetic variants are randomly allocated at conception and remain independent of most environmental and behavioral factors^5,6^. In addition, recent large-scale genome-wide associations studies (GWAS) of circulating metabolic traits^7–11^ and cardiovascular diseases^12–18^ provide valuable resources, offering a strong foundation for MR studies to explore the causal relationships between omega-3 levels and cardiovascular outcomes.

To date, at least 16 MR methods have been developed, with key differences in the selection of IVs, modeling, and techniques to control potential pleiotropy^19,20^. Among these, the inverse variance weighed (IVW) fixed-effects model remains the most widely used method. However, due to its higher susceptibility to type I error, MR-Egger^21^, Weighted-mode^22^, and MR-APSS^23^ models were also incorporated in this study. These new methods have shown robust control of type I errors and improved accuracy in estimating causal effects under diverse conditions in both simulated and real-world data^20,24^.

To ensure a robust causal inference, we integrated data from two large independent GWAS of circulating omega-3^9,25^ with genetic data on 14 cardiovascular and metabolic phenotypes, each involving more than 50,000 participants (**Figure 1**). This study aims to systematically examine the causal relationships between omega-3 levels and cardiovascular risk using large-scale genetic data.

**Figure 1.**
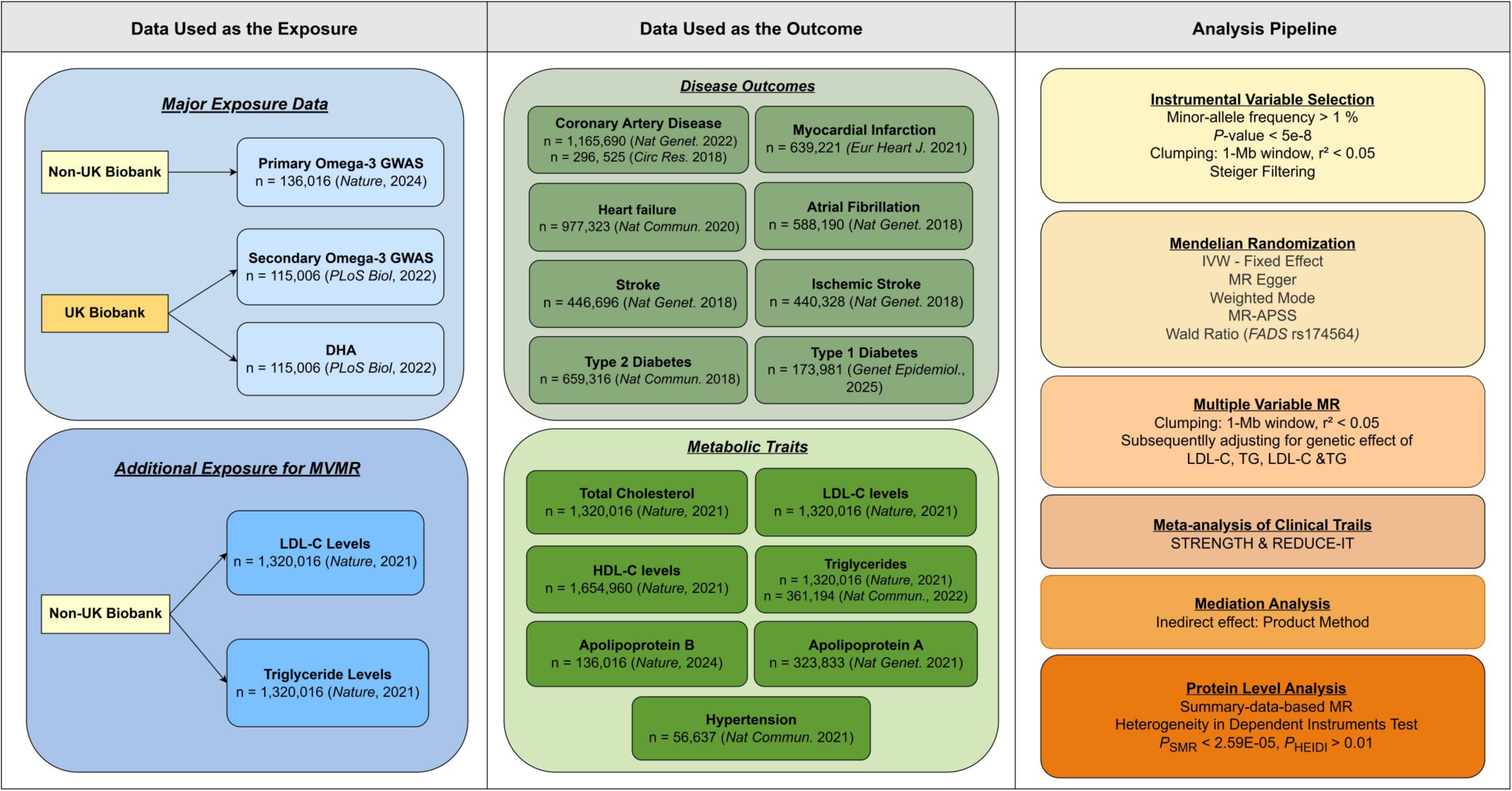
**Flowchart of Study Design.**

## Methods

### Study design and data sources

The full GWAS summary statistics used in this analysis were primarily obtained from the GWAS Catalog^26^, with detailed information provided in **Table S1**. These GWAS were predominantly conducted in European populations. To ensure adequate statistical power, we included only studies with at least 50,000 participants. Patients or the public were not involved in the design, conduct, reporting, or dissemination plans of our research.

We collected two independent GWAS datasets for circulating omega-3 measurement as the exposure (**Figure 1**). The primary exposure dataset was derived from a multi-cohort meta-analysis, involving 136,016 participants^9^, while the secondary exposure dataset was sourced from the UK Biobank, including 115,006 individuals^25^. Notably, there was no sample overlap between these two datasets, strengthening the validity of our findings. DHA measurements from the UK Biobank^25^ were also included as the exposure. Summary statistics for other metabolic traits, including total cholesterol, low-density lipoprotein cholesterol (LDL-C), high-density lipoprotein cholesterol (HDL-C), triglyceride, apolipoprotein B (ApoB), apolipoprotein A (ApoA)^7–10^, were also mainly sourced from the two studies. Most participants in the multi-cohort study were fasted before blood collection^9^, whereas the UK Biobank participants were generally non-fasted, with precise fasting intervals recorded and adjusted for in the analyses^25^. Both studies controlled for age, sex, fasting status, sample type, and relevant covariates, such as body mass index (BMI) and smoking status. These analytical strategies have been well validated and demonstrated in their publications^9,25^.

We also collected GWAS summary statistics for a broad range of cardiovascular outcomes, including coronary artery disease (CAD)^12,14^, myocardial infarction (n = 639,221)^13^, heart failure (n = 977,323)^15^, atrial fibrillation (n = 583,168)^16^, stroke (n = 446.696)^17^, and its ischemic subtype (n = 440328)^17^. Two CAD GWAS datasets were analyzed: (i) a recent European meta-analysis with 181,522 cases and 1,165,690 controls^12^, and (ii) the UK Biobank (34,541 cases and 261,984 controls)^14^. We also incorporated data for hypertension (n = 56,637)^11^ and type 2 diabetes (T2D, n = 659,316)^18^ due to their strong connection to cardiovascular disease, while type 1 diabetes (T1D, n = 173,981)^27^, a type of autoimmune diseases, was included for comparison. Age, sex, principal components and relevant study-specific covariates were adjusted and validated in their publications.

We used the STROBE reporting guideline^28–30^ to draft this manuscript, and the STROBE reporting checklist when editing, included in **Table S7.**

### Instrumental variable selection

Common SNPs (minor-allele frequency > 1 %) associated with omega-3 levels at genome-wide significance level (*P* < 5.00E-08) were selected. Correlated variants were then removed by linkage disequilibrium (LD) clumping (1-Mb window, r² < 0.05) based on the 1,000 Genomes European panel. To mitigate pleiotropic SNPs, we further applied Steiger filtering^31^ discarding SNPs that explained more variance in the outcome than in the exposure, maintaining the correct causal direction.

### Mendelian randomization analyses

To ensure the robustness of our MR results, we employed four well-established MR algorithms, as evaluated in previous studies^19,20^. These included: (i) IVW Fixed-effects^32^, (ii) MR-Egger^21^, (iii) Weighted Mode^22^, and (iv) APSS^33^. In addition, given that the *FADS* variant accounts for a large proportion of variance in omega-3 levels^34^, we performed a single-variant Wald Ratio analysis^35^ using rs174564 or a proxy SNP in strong LD (r^2^ > 0.9). A causal association was accepted when at least three of the five tests yielded statistically significant results with consistent directional effects. The significance threshold for *P* values was adjusted using Bonferroni correction.

To ensure comparability between datasets, we standardized the SNP-effect estimates and their standard errors (SE). Each raw effect (β) was divided by its SE to create a Z-score, which we then rescaled by 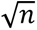 (n = sample size) to produce a unit-free effect (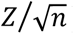). The corresponding SE was set to 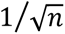. This transformation eliminates differences in measurement units and variance across studies, ensuring that subsequent instrumental-variable analyses are conducted on a consistent scale.

SNP-exposure and SNP-outcome datasets were harmonized using the TwoSampleMR package (version 0.5.11)^36^. IVW Fixed-effects model, MR-Egger, Weighted Mode, and Wald Ratio analyses were also conducted using this package. APSS was implemented with the MRAPSS package (0.0.0.9), which restricted the analysis to SNPs in the HapMap 3 reference panel. The background parameters were estimated by LD score regression^37^, with the LD score calculated from the 1,000 Genomes European panel.

### Multivariable mendelian randomization analysis

Multivariable mendelian randomization (MVMR) analysis was performed using the TwoSampleMR package (version 0.5.11) to estimate the direct association of omega-3 levels with CAD. The models were sequentially adjusted for LDL-C, triglycerides and then for both together. IVs for omega-3 fatty acids, LDL-C, and triglycerides were selected independently based on predefined criteria. LD clumping (1-Mb window, r² < 0.05) was subsequently conducted to ensure instrument independence. Effect alleles were harmonized across both exposures and the outcome datasets.

### Mediation analysis

Mediation analysis^38^ was applied to partition the effect of genetically predicted omega-3 on the CAD into a direct and an indirect effect mediated through LDL-C. The total effect was estimated using the four well established MR algorithms, respectively. Indirect effects were assessed using a two-step MR framework (**Figure 4b**): (1) estimating the effect of omega-3 on LDL-C (β_step1_), and (2) estimating the effect of LDL-C on CAD (β_step2_). The indirect effect was then calculated using the product method as β_step1 ×_ β ^39,40^.

To estimate the SE of the indirect effect, we applied two approaches: (1) the Delta method approximates the variance using the delta rule, yielding a closed-form solution (SE_indirect_ = sqrt(β ^2^ * SE_step2_^2^ + β ^2^ * SE_step1_^2^)^41^ but potentially underestimating variability when coefficient distributions are skewed; (2) the Monte Carlo (MC) method generated empirical confidence intervals by repeatedly sampling^42^, offering greater robustness at the expense of higher computational cost. Both methods were implemented to ensure robustness of our inference.

### Protein quantitative trait loci data analysis

Summary-data-based mendelian randomization (SMR)^43,44^ was applied to identify plasma proteins associated with omega-3 levels. To differentiate true associations from those arising due to LD, we incorporated the heterogeneity in dependent instruments (HEIDI) test^44^. These analyses were conducted using the SMR portal^43^, leveraging protein quantitative trait loci (pQTL) summary statistics obtained from the Fenland study (n = 10,708)^45^. Associations were considered statistically significant if they met the following criteria: (i) the lead SNP in omega-3 GWAS passed the genome-wide significant threshold (*P* < 5.00E-08); (ii) the SMR test passed a Bonferroni-corrected threshold (*P*_SMR_ < 2.59E-05; 0.05/1,927); and (iii) there was no evidence of horizontal pleiotropy due to LD (*P*_HEIDI_ > 0.01). Proteins were considered associated with omega-3 if significant associations were confirmed in both omega-3 GWAS. Gene set enrichment analysis was conducted using ToppGene Suite^46^. Regional plots were generated by R package locuscomparer (Version 1.0.0)^47^.

## Results

### Genetically elevated omega-3 levels increase the risk of cardiovascular outcomes

Summary statistics from a GWAS of omega-3 measure in 136,016 participants^9^ were used as the primary exposure dataset. The causal effects on seven cardiovascular-related outcomes were examined using five MR estimates (**Methods**). A causal association was considered robust when at least three methods showed consistent effect directions and met the Bonferroni-corrected significance threshold (*P* < 0.00625; 0.05/8).

Our analysis showed that genetically elevated omega-3 levels have a significant causal effect on the risk of CAD (*P*_median_ = 2.65E-04), with consistent effect directions across all MR estimates (**Figure 2a**). These results were consistent when using the CAD GWAS exclusively from the UK Biobank (*P*_median_ = 2.87E-07). Similar causal relationships were observed for myocardial infarction (*P*_median_ = 1.03E-07) and heart failure (*P*_median_ = 4.84E-03). We also observed modest effects of omega-3 on the risk of atrial fibrillation (*P*_median_ =0.041), stroke (*P*_median_ = 0.038) and its ischemic subtype (*P*_median_ = 0.035). Although these associations did not meet the Bonferroni-corrected significance threshold, the directions of causal effects were consistent across all five MR estimates. In comparison, no causal effects were observed for T2D (*P*_median_ = 0.139) and T1D (*P*_median_ = 0.297).

**Figure 2.**
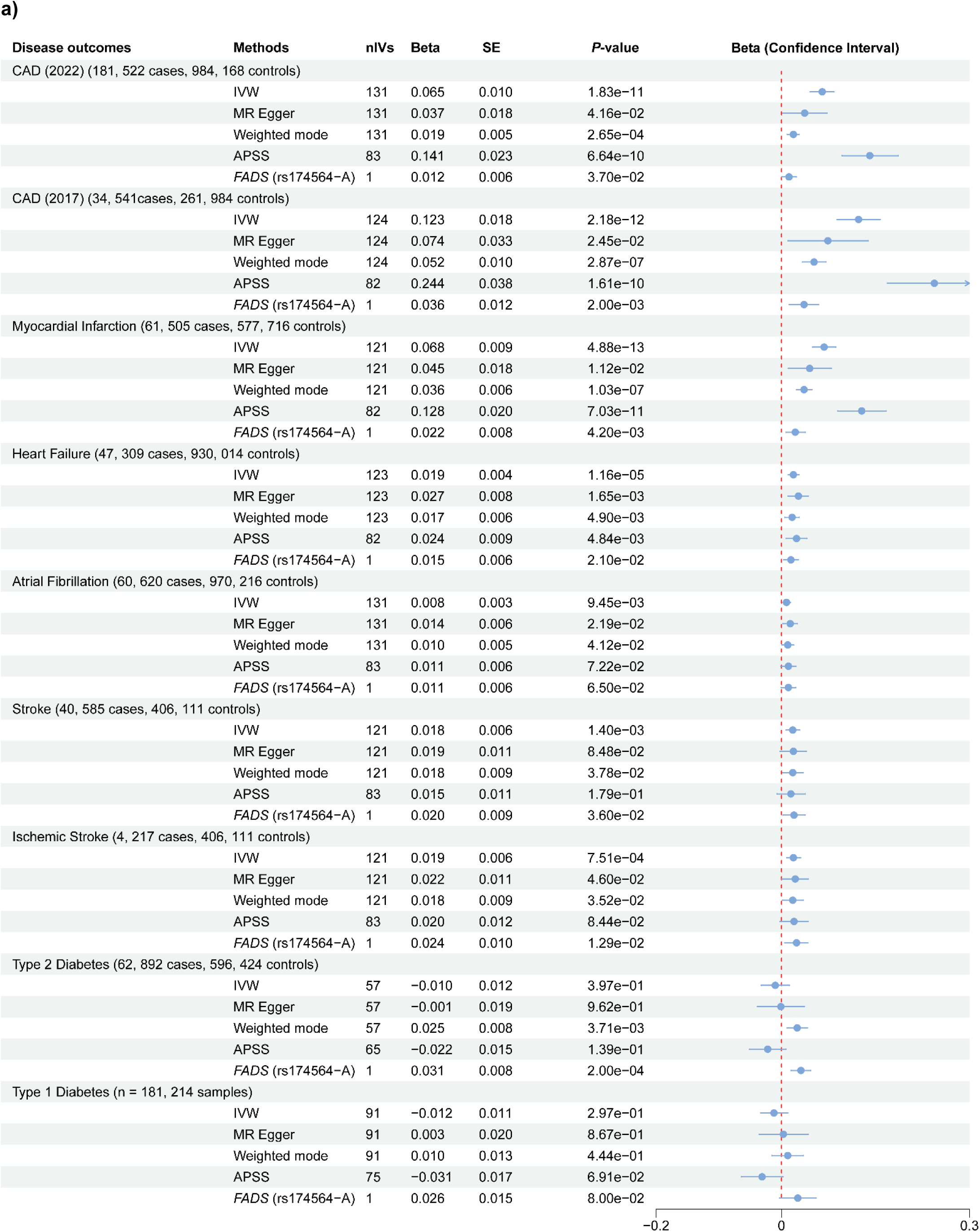
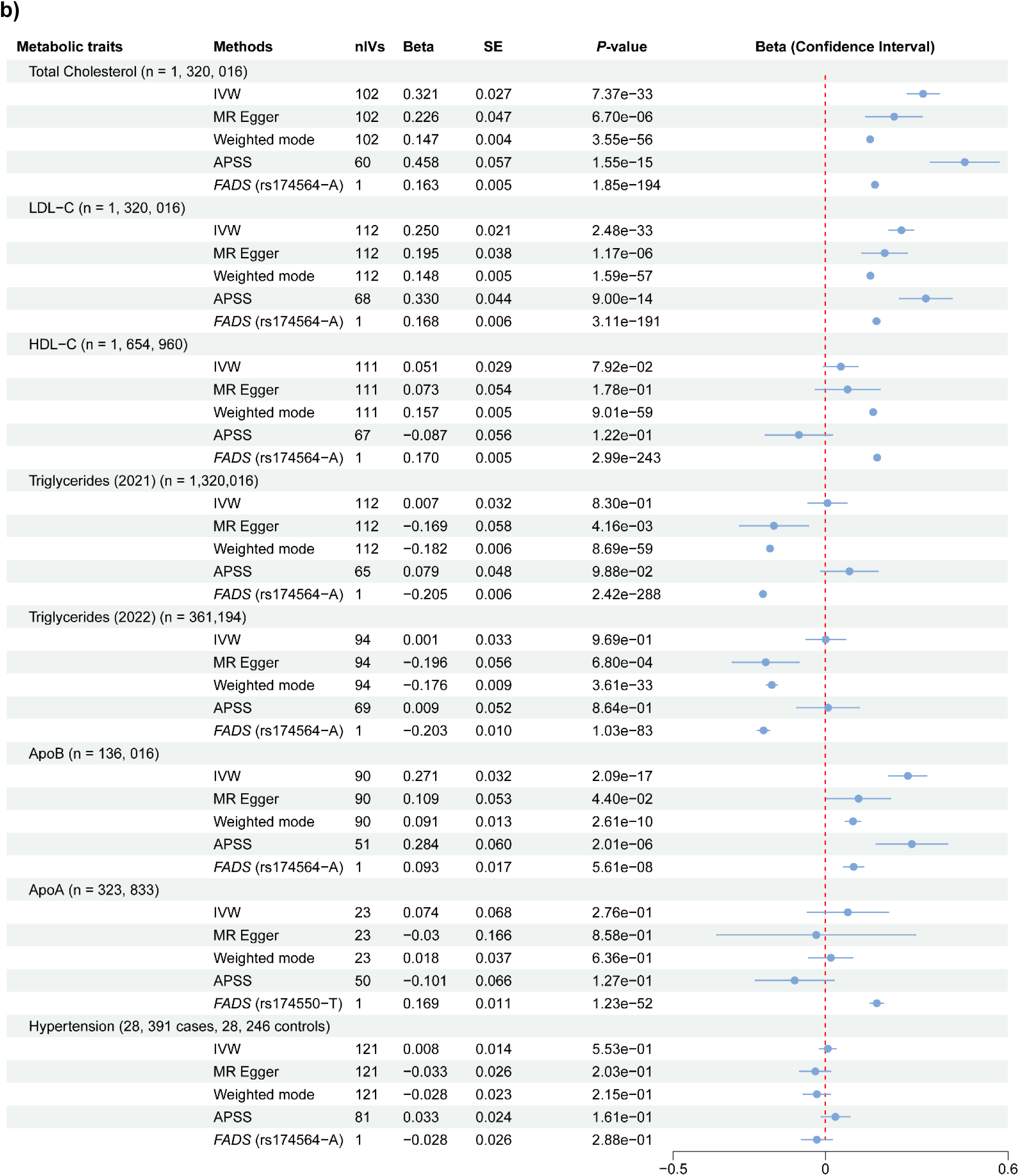
Effect of omega-3 on cardiovascular events (a) and related metabolic traits (b) estimated across five mendelian randomization (MR) methods. nIVs, number of instrumental variables (IVs) used in the analysis. SE, standard error of the effect estimate. The APSS algorithm employs considerably fewer instrumental variables because it restricts analysis to SNPs included in the HapMap 3 reference panel (See Methods).

We repeated the analysis using the secondary omega-3 GWAS from UK Biobank. The results confirmed the causal association of increased omega-3 levels with higher risks of CAD (*P*_median_ = 2.94E-03 and 5.45E-05 for respective datasets), myocardial infarction (*P*_median_ = 1.27E-03) and heart failure (*P*_median_ = 4.60E-03). All five MR estimates pointed in the same direction (**Figure S1a**). The positive association with atrial fibrillation (*P*_median_ = 0.065), stroke (*P*_median_ = 0.140), and ischemic stroke (*P*_median_ = 0.044) still exists, whereas no associations were detected for T2D (*P*_median_ = 0.126) and T1D (*P*_median_ = 0.727). We also repeated the analyses using a GWAS of DHA as the exposure. The results were highly consistent, demonstrating a significant causal effect of elevated DHA levels on the risks of cardiovascular outcomes (**Figure S2**).

We next conducted sensitivity analyses by excluding genetic variants in the *FADS* gene cluster (hg19; chr11:61,067,097–62,134,826), as the fatty acid desaturases encoded in this region directly involve omega-3 metabolism^48^. The causal association with CAD (*P*_median_ = 9.47E-08 and 8.39E-07 for respective datasets), myocardial infarction (*P*_median_ = 5.70E-06) and heart failure (*P*_median_ = 0.016) remained significant and directionally consistent (**Figure 3a**). These findings were consistent in the secondary omega-3 GWAS (**Figure 3b**).

**Figure 3.**
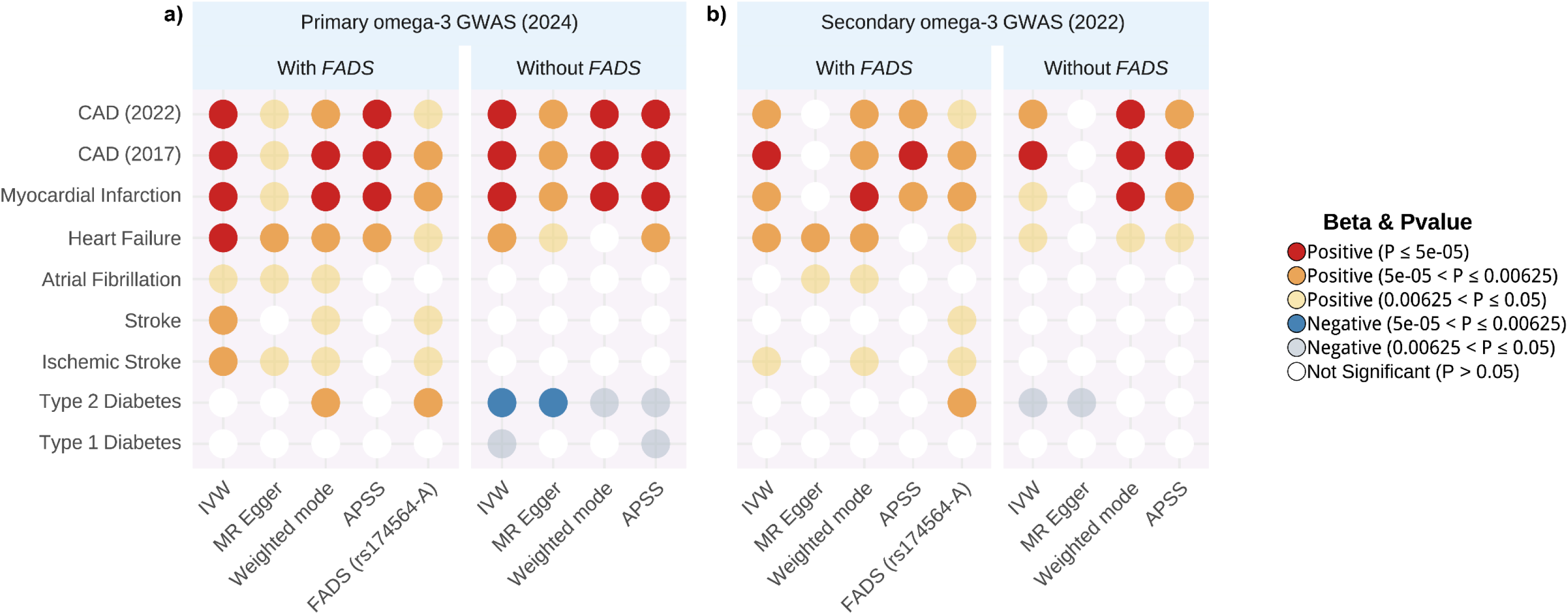
Effects of omega-3 on cardiovascular events with or without inclusion of variants in the FADS gene cluster. (a) Primary omega-3 GWAS derived from a multi-cohort study was used as the exposure. (b) Secondary omega-3 GWAS derived from UK Biobank (2022) was used as the exposure. The *FADS* gene cluster was defined as a region in hg19; chr11:61,067,097–62,134,826. The Bonferroni-adjusted significance threshold was set to be 0.00625 (0.05/8).

**Figure 4.**
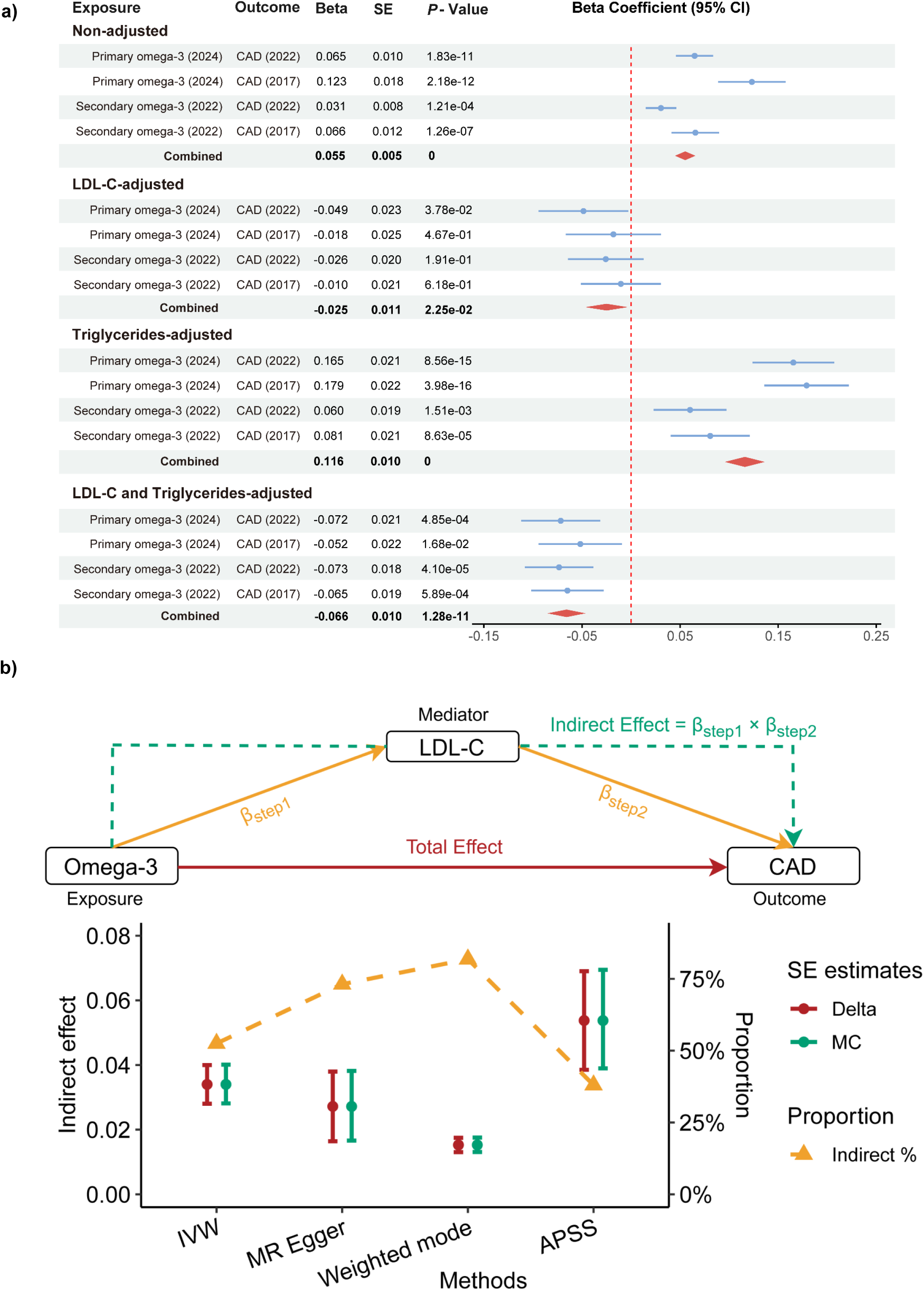
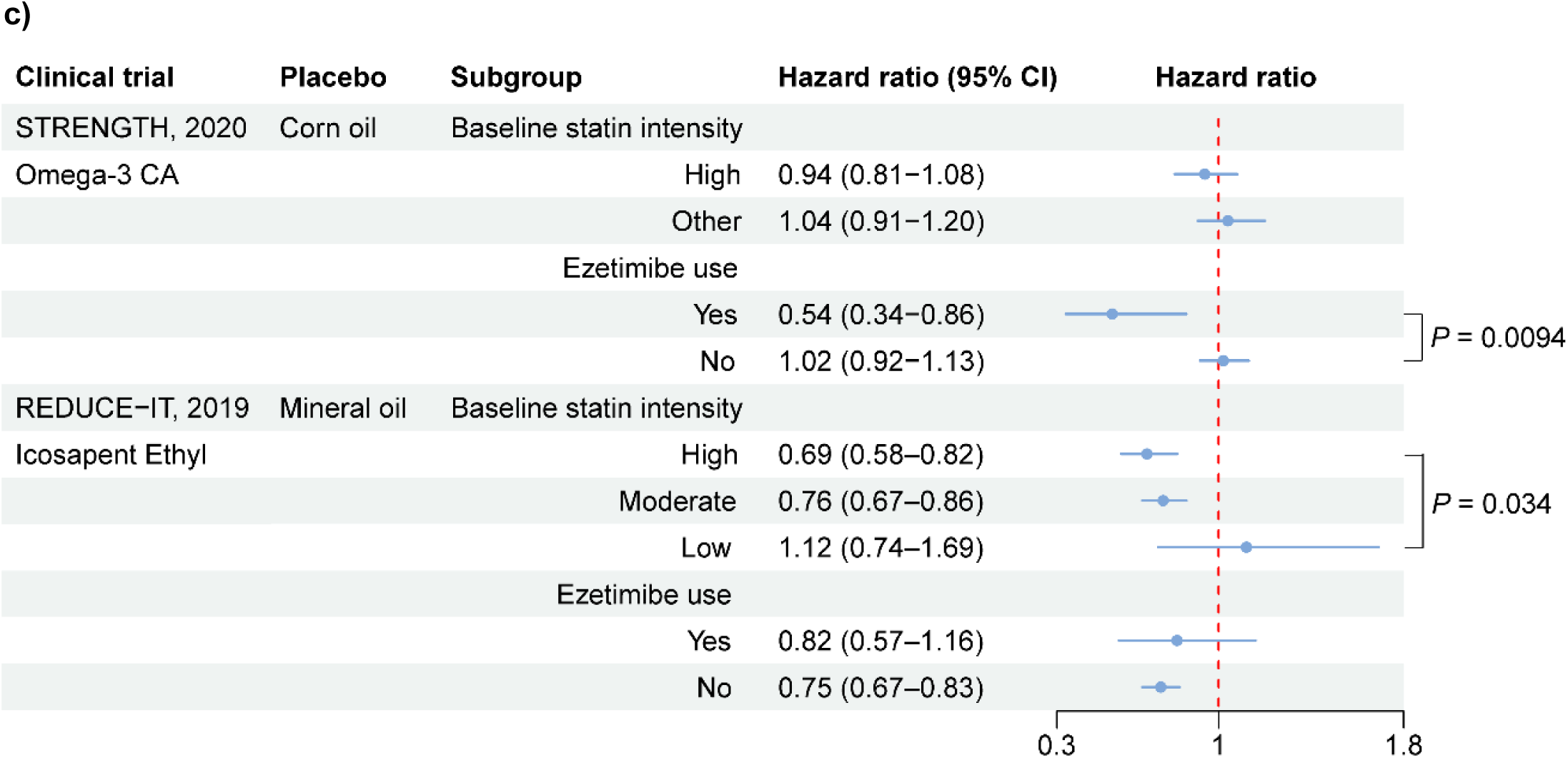
Enhanced Lipid control is essential for omega-3’s cardiovascular protection. a) The direct effect of omega-3 on coronary artery disease by subsequently adjusting for variants associated with LDL-C, triglycerides and both lipid traits. b) Mediation analysis of the effect of omega-3 fatty acids on CAD risk through LDL-C. Red solid line illustrates the total effect of genetically predicted omega-3 on CAD. Step 1 (yellow solid line) represents the effect of omega-3 on LDL-C, and Step 2 (yellow solid line) represents the effect of LDL-C on CAD. The green dashed line indicates the indirect effect mediated by LDL-C. Indirect effect estimates of omega-3 on CAD through LDL-C using 4 MR methods. Error bars represent 95% CIs calculated using the Delta method (red) and Monte Carlo method (green). Yellow triangles represent proportion of the indirect effect relative to the total effect across 4 MR methods. c) Subgroup analyses from the STRENGTH and REDUCE-IT trials showed that the protective effect of omega-3 appeared to be associated with the intensity of LDL-C lowering therapy.

Reverse MR analyses were also conducted using genetic variants associated with cardiovascular outcomes as IVs. No evidence of reverse causal associations was observed (**Figure S3**), suggesting a unidirectional relationship in which omega-3 levels influence cardiovascular outcomes. Taken together, our results suggest that elevated omega-3 levels may increase cardiovascular risk, particularly CAD, myocardial infarction, and heart failure.

### Omega-3 elevates LDL-C while lowering triglycerides

We further examined the causal influence of omega-3 fatty acids on related metabolic traits. Similarly, a causal association was considered robust if at least three methods showed consistent directions and met the Bonferroni-corrected significance threshold (*P* < 0.0071; 0.05/7).

Our analyses indicated that genetically elevated omega-3 levels significantly increased total cholesterol (*P*_median_ = 7.40E-33), especially LDL-C (*P*_median_ = 2.50E-33). These results were highly consistent across the five MR methods (**Figure 2b**). In contrast, no significant effect was observed for HDL-C (*P*_median_ = 0.079) and hypertension (*P*_median_ = 0.215). This pattern extended to key apolipoproteins, with a significant effect found on ApoB (*P*_median_ = 5.60E-08), the key component of LDL particles, while no association was found for ApoA (*P*_median_ = 0.276), the key component of HDL particles. The causal effects on LDL-C and ApoB were also replicated using another independent omega-3 GWAS from UK Biobank (**Figure S1b**). Sensitivity analyses were also performed by excluding genetic variants within the *FADS* gene cluster. Strong positive causal associations of omega-3 with LDL-C and ApoB levels were still observed (**Figure S4**).

To address potential influence of horizontal pleiotropy between omega-3 and LDL-C mentioned in previous studies^49^, we adopted more stringent criteria for selecting IVs. Only genetic variants associated with omega-3 levels (*P* < 5.00E-08) but not associated with LDL-C levels (*P* > 0.05), were included in the MR analyses. Despite the reduced statistical power, a positive causal association between omega-3 and LDL-C remained evident *(P* < 0.019; **Figure S5**).

In addition to LDL-C, our analyses demonstrated that genetically elevated omega-3 levels significantly lowered triglycerides (*P*_median_ = 4.16E-03). This result was also confirmed by another triglyceride GWAS data^8^ (*P*_median_ = 6.80E-04; **Figure 2b**). A consistent direction of effect on triglyceride lowering was also observed when utilizing the omega-3 GWAS derived from UK Biobank as the exposure (**Figure S1b**). These findings align with the American Heart Association (AHA)’s recommendation of omega-3 for reducing triglycerides^50^

However, exclusion of genetic variants within the *FADS* gene cluster reversed this association in a positive direction. This result was further validated using the secondary omega-3 GWAS data (**Figure S4**), highlighting the complex role of omega-3 in triglyceride reduction. Specifically, the triglyceride-lowering effect could be primarily mediated through fatty acid desaturase activity, while omega-3-associated variants outside the *FADS* cluster were linked to increased triglyceride levels. These findings suggest that the effectiveness of omega-3 supplementation in reducing triglycerides may depend on an individual’s fatty acid desaturase activity.

To validate these findings, we revisited data from previous clinical trials. To minimize the influence of cholesterol-lowering drugs, we first analyzed data from the Comparing EPA to DHA trial (ComparED; NCT01810003)^51^, which assessed changes in blood lipids following EPA or DHA intake among adults without a history of cardiovascular disease^51^. Compared to corn oil, both EPA and DHA significantly increased LDL-C levels while reducing triglycerides. This effect was markedly stronger with DHA, demonstrating a mean LDL-C increase of 0.16 mmol/L (*P* < 0.0001) and a mean triglyceride reduction of 0.25 mmol/L (*P* < 0.0001) over a 10-week follow-up^51^.

Next, we revisited data from the STRENGTH trial^4^. Although all participants were on statin therapy, those in the omega-3 CA group still showed a significant increase in LDL-C compared with the corn oil group (geometric mean ratio = 1.03; *P* < 0.001) and a substantial reduction in triglyceride levels (geometric mean ratio = 0.82; *P* < 0.001)^4^.

We also investigated data from the REDUCE-IT trial^3^. Due to the use of mineral oil as the control, between-group comparisons were not considered. As all participants were receiving statin therapy, LDL-C were expected to be reduced^52^. However, within the icosapent ethyl group, LDL-C levels increased by 3.1 % from the baseline (*P* < 0.001). Triglycerides decreased by 18.3 % (*P* < 0.001) following 12 months of treatment. Collectively, findings from previous clinical trials supported our MR results that omega-3 fatty acids can elevate LDL-C levels while lowering triglycerides.

### The protective effect of omega-3 on CAD depends on lipid control

To evaluate the impact of LDL-C, we estimated the effect of omega-3 on cardiovascular outcome by controlling LDL-C-associated variants using MVMR algorithm (**Methods**). To maximize statistical power, CAD was selected as the outcome due to its largest sample size and the availability of additional CAD dataset for replication. Our results showed that, after adjusting for LDL-C’s genetic contribution, the effect of omega-3 on CAD became protective. The association remained consistent when replicated using the secondary omega-3 GWAS or an alternative CAD GWAS, yielding a combined effect estimate of approximately −0.025 (*P*_combined_ = 0.0225) (**Figure 4a** and **Table S2**). In contrast, adjusting for the genetic variants associated with triglycerides didn’t change the direction of the associations (**Figure 4a** and **Table S3**). However, we found that adjusting for the genetic effects of both LDL-C and triglycerides further strengthened omega-3’s protective association with CAD (β_combined_ = −0.066, *P*_combined_ = 1.28E-11; **Figure 4a** and **Table S4**). These findings highlight the critical role of lipid control, particularly LDL-C, in optimizing the cardioprotective effects of omega-3.

To further evaluate the role of LDL-C in mediating the effect of omega-3 fatty acids on CAD risk, we utilized mediation analysis to decompose the total effect into direct and indirect components (**Methods**). The indirect effect on CAD through LDL-C was significant (β_median_ = 0.031; *P* _median_ = 2.40E-12; **Figure 4b**), explaining approximately 38.0–81.9% of the total effect across four MR estimates (**Figure 4b** and **Table S5**). These results remained robust when replicated using an alternative CAD GWAS (**Figure S6** and **Table S5**). These findings suggest LDL-C as a major mediator of the association between elevated omega-3 and CAD risk and highlight the potential importance of targeting LDL-C reduction in this pathway.

To validate the significance of LDL-C management in optimizing omega-3 benefits, we reanalyzed data from the STRENGTH and REDUCE-IT trials, both of which evaluated the effect of high-dose omega-3 on cardiovascular events^3,4^. Subgroup analyses support our findings (**Figure 4c)**. In STRENGTH, compared to corn oil, omega-3 CA appeared more effective in participants receiving high-intensity statin therapy (Hazard Ratio [HR] = 0.94) than in other participants (HR = 1.04), though this difference did not reach statistical significance (*P* = 0.28). Notably, in participants receiving combined statin and ezetimibe therapy, a regimen that achieves greater LDL-C reduction, omega-3 CA significantly lowered cardiovascular risk compared to corn oil (HR = 0.54, 95% Confidence Interval [CI]: 0.34-0.86; *P* = 0.0094)^53^.

A similar trend was observed in the REDUCE-IT trial, where icosapent ethyl conferred a significant cardiovascular benefit over mineral oil among participants receiving high-intensity statin therapy (HR = 0.69, 95% CI: 0.58–0.82; *P* = 2.60E-05). In contrast, no benefit was seen in the low-intensity statin group (HR = 1.12, 95% CI: 0.74–1.69; *P* = 0.59). The difference between the two groups was significant (Cochran’s Q, *P* = 0.034), indicating that patients on high-intensity statin therapy gained substantially more from icosapent ethyl than those on low-intensity therapy. In the smaller subgroup receiving both a statin and ezetimibe, icosapent ethyl also suggested a protective effect (HR = 0.82; 95% CI, 0.57–1.16), although this was not significantly different from the statin-only group. This may be due to the limited sample size and the fact that 93% of participants were already on at least moderate-intensity statin therapy^3^. Taken toghether, our findings suggest the importance of lowering LDL to optimize the cardioprotective effects of omega-3. However, the potential for enhanced benefits through concurrent management of triglycerides and LDL-C requires further validation.

### Plasma proteins associated with omega-3 levels

Circulating proteins are often used as biomarkers or targets for pharmacological intervention^45^. We incorporated pQTL data from the Fenland study (n = 10,708) to detect plasma proteins associated with omega-3 levels. Using SMR analysis combined with the HEIDI test (**Methods**), we identified 9 candidate proteins that passed the Bonferroni-corrected threshold (*P*_SMR_ < 2.59E-05; 0.05/1,927). Of these, five were further replicated in the secondary omega-3 GWAS data (**Figure 5**), including Glucokinase Regulator (GCKR), Angiopoietin Like 3 (ANGPTL3), Proprotein Convertase Subtilisin/Kexin Type 9 (PCSK9), T Cell Immunoglobulin And Mucin Domain Containing 4 (TIMD4), and ApoB (**Table S6**). Regional plots revealed that genetic variants linked to the expression of these proteins well colocalized with signals for omega-3 levels (**Figure S7 - S8**), highlighting strong associations between these circulating proteins and omega-3 metabolism.

**Figure 5.**
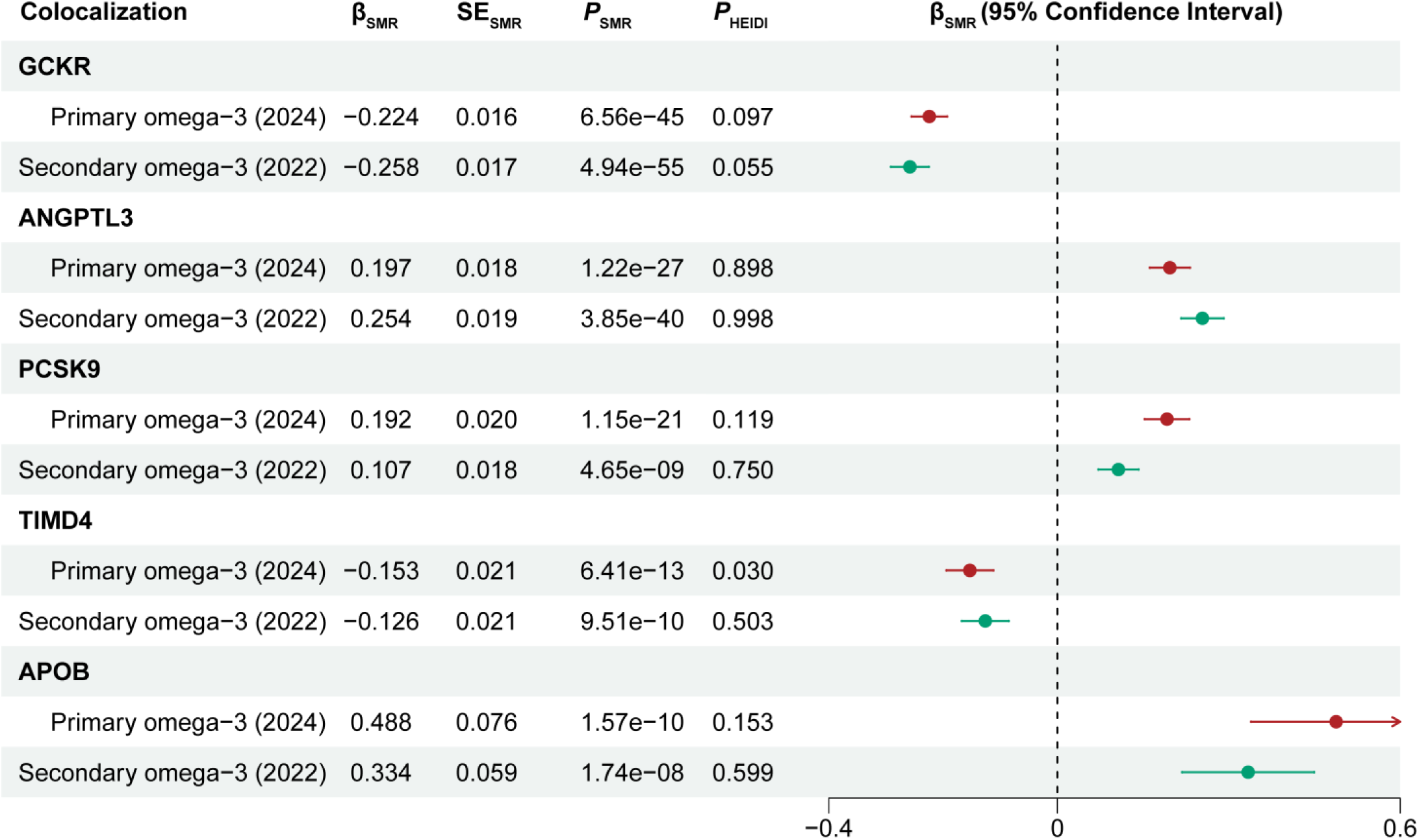
Colocalization of protein QTLs (pQTLs) with genetic loci associated with omega-3 levels. Red dots show the results derived from the primary omega-3 GWAS, and green dots represent the results based on the secondary omega-3 GWAS. SE stands for Standard error.

Gene set enrichment analysis demonstrated that these proteins were highly enriched in pathways related to lipid homeostasis (GO:0055088; FDR = 1.91E-04) and triglyceride metabolic process (GO:0006641; FDR = 2.04E-04). Of these, ANGPTL3 (β_SMR_ = 0.197, *P*_SMR_ = 1.22E-27 and *P*_HEIDI_ = 0.898), PCSK9 (β_SMR_ = 0.192, *P*_smr_ = 1.15E-21 and *P*_HEIDI_ = 0.119), and ApoB (β_SMR_ = 0.488, *P*_smr_ = 1.57E-10 and *P*_HEIDI_ = 0.153) demonstrated a positive correlation with omega-3 levels (**Figure 5**). Their upregulations have been found linked to increased LDL-C levels and a higher risk of cardiovascular events^54–56^. These genetic findings suggest a potential mechanism whereby higher omega-3 levels are positively associated with proteins known to increase LDL-C (PCSK9, ANGPTL3, ApoB), which could partly explain their potential link to increased cardiovascular risk. However, this hypothesis requires further mechanistic studies and clinical validation.

Notably, PCSK9 and ANGPTL3 are established targets for FDA-approved drugs. Evolocumab, a PCSK9 inhibitor, has been shown to reduce LDL-C levels by 59% compared to placebo at 48 weeks^57^, while Evinacumab, an ANGPTL3 inhibitor, achieves a 49% reduction in LDL-C and approximately 50% reductions in triglyceride levels relative to placebo at 24 weeks in patients with familial hypercholesterolemia^58^. In light of the MVMR and mediation analyses, omega-3 supplementation could be a valuable complement to PCSK9-targeted and ANGPTL3-targeted therapies. These hypotheses warrant further investigation.

## Discussion

Using large-scale genetic data, we showed that genetically elevated omega-3 levels were associated with increased cardiovascular risk, particularly CAD, myocardial infarction, and heart failure. These results were consistent across independent omega-3 GWAS datasets and different MR algorithms. Additional sensitivity analyses also supported the positive causal relationship between higher omega-3 levels and increased cardiovascular risk. Although prior studies, such as STRENGTH and REDUCE-IT, reported neutral or protective effects, our results don’t violate their findings, as these studies were conducted in patients receiving LDL-lowering therapy. Our MVMR and mediation analyses showed that effective LDL control is essential for omega-3 to confer cardiovascular benefits, providing a plausible explanation for the differing results between the disease population and the general population.

Notably, a recent prospective cohort study showed that regular use of fish oil supplements was linked to a higher risk of first-time heart disease in individuals without prior cardiovascular conditions, but slowed disease progression in those with existing cardiovascular disease^59^. The differences may also reflect the role of LDL control in mediating the protective effect of omega-3, as individuals without cardiovascular conditions often use omega-3 supplementation without concurrent LDL-lowering therapy. Emerging evidence^60^ also suggests that LDL-C may act as a mediating factor between increased fatty acids biosynthesis and higher cardiovascular risk. Consistent with these findings, further analyses from the STRENGTH^4^ and REDUCE-IT^3^ trials indicate that the protective effect of omega-3 on CAD is more pronounced with intensive LDL-C-lowering therapy. Together, these findings highlight the need for optimal lipid management, particularly LDL-C, when using omega-3 supplementation to achieve cardiovascular protection.

Analysis of metabolic traits revealed that elevated omega-3 levels were significantly linked to increased LDL-C, ApoB, and total cholesterol levels. Circulating proteins known to increase LDL-C, such as PCSK9, ANGPTL3, ApoB, were also found positively associated with omega-3 levels. These findings well explain the positive association observed between omega-3 and cardiovascular risk. However, mechanistic studies are needed to clarify the underlying pathways. Omega-3 also showed a triglyceride-lowering effect, though this benefit appeared to be mainly driven by the *FADS* gene cluster. Previous clinical trials, including ComparED, STRENGTH and REDUCE-IT, also support these findings.

Additionally, PCSK9 and ANGPTL3 are established targets of approved lipid-lowering drugs^61,62^. Their positive associations with omega-3 levels raise the possibility that omega-3 supplementation may interact with these pathways. Further clinical studies are needed to assess whether combining omega-3 with therapies targeting PCSK9 or ANGPTL3 could enhance cardiovascular benefits.

We acknowledge that MR methods rely on strong assumptions^23,63^. The selection of valid IVs is crucial for producing reliable results. We restricted the analyses to GWAS with large sample size (n > 50,000) and applied Steiger filtering to exclude potential pleiotropy. Multiple well-established MR methods were also utilized in combination to further enhance the reliability of the findings. Despite these, completely ruling out horizontal pleiotropy and ensuring sufficient statistical power remain challenging. In addition, some MR analyses were conducted based on a one-sample test, where both exposure and outcomes were collected from the same source, such as the UK Biobank. This approach can minimize differences in population ancestry and composition, but it is more susceptible to overfitting and weak instrument bias^64^.

In summary, this study suggests that the cardiovascular benefits of omega-3 fatty acids may depend on effective lipid management, particularly LDL-C control. We hypothesize that combining omega-3 supplementation with effective lipid-lowering therapy may help maximize cardiovascular benefit, a concept that warrants further clinical investigation.

## Conclusion

Our results suggest that the cardiovascular benefits of omega-3 fatty acids are context-dependent, showing protective effects when lipids, particularly LDL-C, are well controlled. We hypothesize that combining omega-3 supplementation with effective lipid-lowering therapy may help optimize cardiovascular benefit, a concept that warrants further clinical investigation.

## List of abbreviations

CAD: Coronary artery disease
MVMR: Multivariable Mendelian randomization
SMR: Summary-data-based Mendelian randomization
PUFAs: Polyunsaturated fatty acids
RCTs: Randomized clinical trials
MR: Mendelian randomization
IVs: Instrumental variables
GWAS: Genome-wide association studies
IVW: Inverse variance weighted
LDL-C: Low-density lipoprotein cholesterol
HDL-C: High-density lipoprotein cholesterol
ApoB: Apolipoprotein B
ApoA: Apolipoprotein A
BMI: Body mass index
T2D: Type 2 diabetes
T1D: Type 1 diabetes
LD: Linkage disequilibrium
SE: Standard errors
HEIDI: Heterogeneity in dependent instruments test
pQTL: Protein quantitative trait loci

## Data Availability

https://www.ebi.ac.uk/gwas/studies/GCST90301959
https://www.ebi.ac.uk/gwas/studies/GCST90092931
https://www.ebi.ac.uk/gwas/studies/GCST90092816
https://www.ebi.ac.uk/gwas/studies/GCST90132314
https://www.ebi.ac.uk/gwas/studies/GCST005194
https://www.ebi.ac.uk/gwas/studies/GCST011365
https://www.ebi.ac.uk/gwas/studies/GCST009541
https://www.ebi.ac.uk/gwas/studies/GCST006061
https://www.ebi.ac.uk/gwas/studies/GCST006906
https://www.ebi.ac.uk/gwas/studies/GCST006908
https://cnsgenomics.com/content/data
https://www.ebi.ac.uk/gwas/studies/GCST90013791
https://www.ebi.ac.uk/gwas/studies/GCST90239676
https://www.ebi.ac.uk/gwas/studies/GCST90239658
https://www.ebi.ac.uk/gwas/studies/GCST90239649
https://www.ebi.ac.uk/gwas/studies/GCST90239664
https://www.ebi.ac.uk/gwas/studies/GCST90179149
https://www.ebi.ac.uk/gwas/studies/GCST90301946
https://www.ebi.ac.uk/gwas/studies/GCST90019495
https://www.ebi.ac.uk/gwas/studies/GCST90086092

## Acknowledgments

We gratefully acknowledge funding supports from the Shenzhen-Hong Kong Jointly Funded Project (Category A; SGDX20230116093201002) and the Stability Support for Higher Education from Shenzhen Science and Technology Program, the Guangdong Natural Science Foundation Youth Enhancement Project (2024A1515030287), 1+1+1 CUHK-CUHK(SZ)-GDSTC Joint Collaboration Fund (GRDP2025-056 and YSP05) and the National Natural Science Foundation of China (82471825). We also extend our thanks to the faculty development grant (Project number: H800005) from the Hang Seng University of Hong Kong and the Warshel Institute for Computational Biology and their funding support from Shenzhen City and Longgang District (LGKCSDPT2024001). The authors thank the anonymous reviewers for their constructive comments and valuable suggestions that helped to improve the quality of the manuscript.

## Author contributions

Y.-F. Wang conceived the study. J.-H. Yu and C.-J. Li took the lead in data analysis. J.-H. Yu, C.-J. Z. Liu, Li, Y.-D. Chen, Y. Lei, D.-Y. Li, W. Ma, Y. Wang, Y. Yang, and R. Wang collected GWAS summary data. J.-H. Yu, C.-J. Li, Y. Wang, Y. Yang, R. Wang, and Y.-F. Wang contributed to the interpretation of the findings. J.-H. Yu, C.-J. Li and Y.-F. Wang wrote the manuscript. All authors read and contributed to the manuscript.

## Data availability

See **Table S1**.

## Declaration of generative AI

We cautiously used ChatGPT 4.1 for linguistic and grammatical refinement to improve the readability during the proofreading. After using this tool, the authors reviewed and edited the content as needed and took full responsibility for the content of the publication.

