## Supplementary material for "Genetic Epidemiological Evidence Indicates the Role of Lipid Control in Omega - 3 – Mediated Cardiovascular Protection": Figure S1-S8

a)

| Disease outcomes | Methods | nIVs | Beta | SE | *P*-value |
|---|---|---|---|---|---|
| CAD (2022) (181, 522 cases, 984, 168 controls) | | | | | |
| | IVW | 159 | 0.031 | 0.008 | 1.21e−04 |
| | MR Egger | 159 | 0.013 | 0.015 | 3.92e−01 |
| | Weighted mode | 159 | 0.014 | 0.005 | 2.94e−03 |
| | APSS | 86 | 0.082 | 0.023 | 5.02e−04 |
| | *FADS* (rs174564−A) | 1 | 0.008 | 0.004 | 3.70e−02 |
| CAD (2017) (34, 541cases, 261, 984 controls) | | | | | |
| | IVW | 132 | 0.066 | 0.012 | 1.26e−07 |
| | MR Egger | 132 | 0.023 | 0.023 | 3.18e−01 |
| | Weighted mode | 132 | 0.035 | 0.008 | 5.45e−05 |
| | APSS | 83 | 0.146 | 0.027 | 1.01e−07 |
| | *FADS* (rs174564−A) | 1 | 0.024 | 0.008 | 2.00e−03 |
| Myocardial Infarction (61, 505 cases, 577, 716 controls) | | | | | |
| | IVW | 140 | 0.031 | 0.010 | 1.27e−03 |
| | MR Egger | 140 | 0.019 | 0.018 | 2.75e−01 |
| | Weighted mode | 140 | 0.023 | 0.005 | 2.40e−05 |
| | APSS | 85 | 0.079 | 0.020 | 8.19e−05 |
| | *FADS* (rs174564−A) | 1 | 0.015 | 0.005 | 4.20e−03 |
| Heart Failure (47, 309 cases, 930, 014 controls) | | | | | |
| | IVW | 138 | 0.014 | 0.004 | 6.78e−04 |
| | MR Egger | 138 | 0.022 | 0.008 | 4.60e−03 |
| | Weighted mode | 138 | 0.013 | 0.004 | 1.85e−03 |
| | APSS | 86 | 0.014 | 0.008 | 8.56e−02 |
| | *FADS* (rs174564−A) | 1 | 0.010 | 0.004 | 2.10e−02 |
| Atrial Fibrillation (60, 620 cases, 970, 216 controls) | | | | | |
| | IVW | 158 | 0.003 | 0.004 | 4.06e−01 |
| | MR Egger | 158 | 0.013 | 0.007 | 4.40e−02 |
| | Weighted mode | 158 | 0.008 | 0.004 | 4.46e−02 |
| | APSS | 86 | −0.01 | 0.008 | 2.14e−01 |
| | *FADS* (rs174564−A) | 1 | 0.008 | 0.004 | 6.50e−02 |
| Stroke (40, 585 cases, 406, 111 controls) | | | | | |
| | IVW | 140 | 0.008 | 0.005 | 9.15e−02 |
| | MR Egger | 140 | 0.011 | 0.009 | 1.96e−01 |
| | Weighted mode | 140 | 0.010 | 0.007 | 1.40e−01 |
| | APSS | 86 | 0.003 | 0.010 | 7.31e−01 |
| | *FADS* (rs174564−A) | 1 | 0.013 | 0.006 | 3.60e−02 |
| Ischemic Stroke (34, 217 cases, 406, 111 controls) | | | | | |
| | IVW | 140 | 0.009 | 0.004 | 3.52e−02 |
| | MR Egger | 140 | 0.015 | 0.008 | 6.88e−02 |
| | Weighted mode | 140 | 0.013 | 0.007 | 4.43e−02 |
| | APSS | 86 | 0.005 | 0.009 | 5.94e−01 |
| | *FADS* (rs174564−A) | 1 | 0.016 | 0.006 | 1.29e−02 |
| Type 2 Diabetes (62, 892 cases, 596, 424 controls) | | | | | |
| | IVW | 49 | −0.02 | 0.013 | 1.26e−01 |
| | MR Egger | 49 | −0.044 | 0.025 | 8.53e−02 |
| | Weighted mode | 49 | 0.006 | 0.010 | 5.62e−01 |
| | APSS | 68 | −0.016 | 0.013 | 1.99e−01 |
| | *FADS* (rs174564−A) | 1 | 0.020 | 0.005 | 2.00e−04 |
| Type 1 Diabetes (n = 181, 214 samples) | | | | | |
| | IVW | 85 | 0.003 | 0.009 | 7.27e−01 |
| | MR Egger | 85 | −0.002 | 0.017 | 8.91e−01 |
| | Weighted mode | 85 | 0.009 | 0.012 | 4.22e−01 |
| | APSS | 75 | 0.003 | 0.017 | 8.59e−01 |
| | *FADS* (rs174564−A) | 1 | 0.017 | 0.010 | 8.00e−02 |

Beta (Confidence Interval)

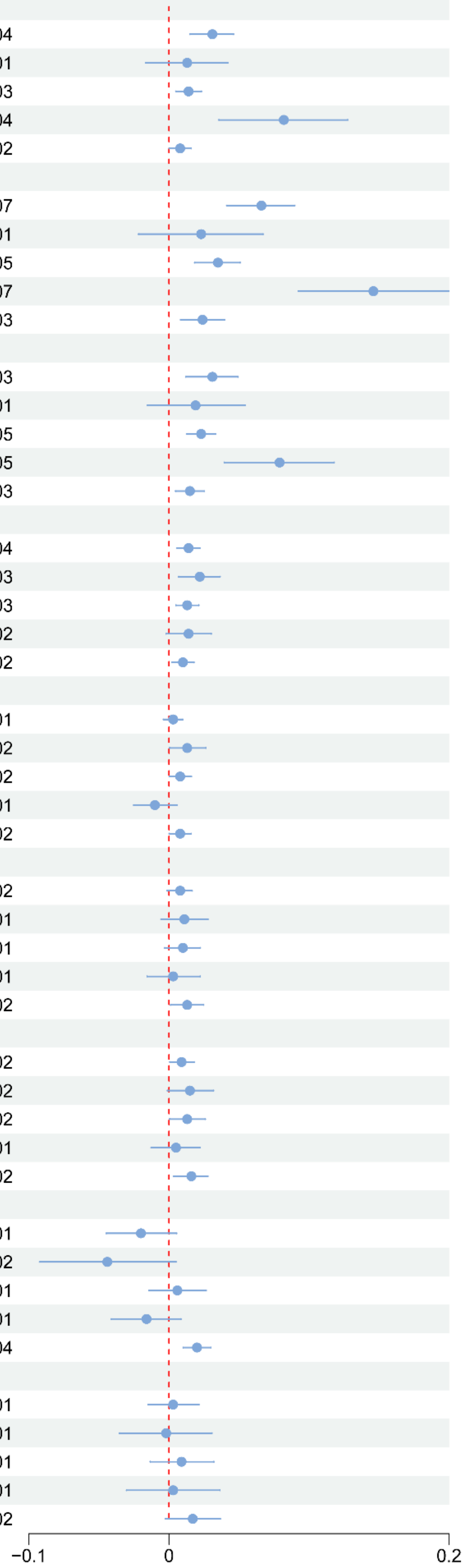

**b)**

| Metabolic traits | Methods | nIVs | Beta | SE | *P*-value |
|---|---|---|---|---|---|
| Total Cholesterol (n = 1, 320, 016) | | | | | |
| | IVW | 151 | 0.253 | 0.021 | 6.76e−34 |
| | MR Egger | 151 | 0.213 | 0.039 | 2.27e−07 |
| | Weighted mode | 151 | 0.105 | 0.003 | 3.33e−67 |
| | APSS | 72 | 0.345 | 0.048 | 7.71e−13 |
| | *FADS* (rs174564−A) | 1 | 0.108 | 0.004 | 1.85e−194 |
| LDL−C (n = 1, 320, 016) | | | | | |
| | IVW | 153 | 0.169 | 0.014 | 1.84e−31 |
| | MR Egger | 153 | 0.155 | 0.027 | 7.38e−08 |
| | Weighted mode | 153 | 0.105 | 0.003 | 1.12e−66 |
| | APSS | 77 | 0.214 | 0.04 | 6.30e−08 |
| | *FADS* (rs174564−A) | 1 | 0.11 | 0.004 | 3.11e−191 |
| HDL−C (n = 1, 654, 960) | | | | | |
| | IVW | 150 | 0.136 | 0.027 | 4.41e−07 |
| | MR Egger | 150 | 0.138 | 0.051 | 7.81e−03 |
| | Weighted mode | 150 | 0.103 | 0.003 | 8.24e−65 |
| | APSS | 75 | 0.082 | 0.057 | 1.51e−01 |
| | *FADS* (rs174564−A) | 1 | 0.112 | 0.003 | 2.99e−243 |
| Triglyceride (2021) (n = 1, 320, 016) | | | | | |
| | IVW | 147 | 0.055 | 0.026 | 3.18e−02 |
| | MR Egger | 147 | −0.067 | 0.047 | 1.62e−01 |
| | Weighted mode | 147 | −0.128 | 0.003 | 6.09e−77 |
| | APSS | 65 | 0.177 | 0.05 | 3.77e−04 |
| | *FADS* (rs174564−A) | 1 | −0.135 | 0.004 | 2.42e−288 |
| Triglyceride (2022) (n = 361, 194) | | | | | |
| | IVW | 103 | 0.026 | 0.03 | 3.79e−01 |
| | MR Egger | 103 | −0.051 | 0.054 | 3.48e−01 |
| | Weighted mode | 103 | −0.129 | 0.007 | 7.64e−34 |
| | APSS | 65 | 0.121 | 0.047 | 9.73e−03 |
| | *FADS* (rs174564−A) | 1 | −0.134 | 0.007 | 1.03e−83 |
| ApoB (n = 136, 016) | | | | | |
| | IVW | 123 | 0.231 | 0.025 | 2.26e−20 |
| | MR Egger | 123 | 0.163 | 0.047 | 6.45e−04 |
| | Weighted mode | 123 | 0.061 | 0.01 | 2.01e−08 |
| | APSS | 67 | 0.35 | 0.053 | 2.95e−11 |
| | *FADS* (rs174564−A) | 1 | 0.061 | 0.011 | 5.61e−08 |
| ApoA (n = 323, 833) | | | | | |
| | IVW | 22 | 0.116 | 0.054 | 3.26e−02 |
| | MR Egger | 22 | 0.012 | 0.089 | 8.92e−01 |
| | Weighted mode | 22 | 0.071 | 0.013 | 1.49e−05 |
| | APSS | 46 | 0.047 | 0.061 | 4.39e−01 |
| | *FADS* (rs174550−T) | 1 | 0.112 | 0.007 | 1.23e−52 |
| Hypertension (28, 391 cases, 28, 246 controls) | | | | | |
| | IVW | 143 | 0.003 | 0.011 | 8.08e−01 |
| | MR Egger | 143 | −0.011 | 0.021 | 5.86e−01 |
| | Weighted mode | 143 | −0.018 | 0.019 | 3.56e−01 |
| | APSS | 82 | 0.036 | 0.025 | 1.49e−01 |
| | *FADS* (rs174564−A) | 1 | −0.019 | 0.017 | 2.88e−01 |

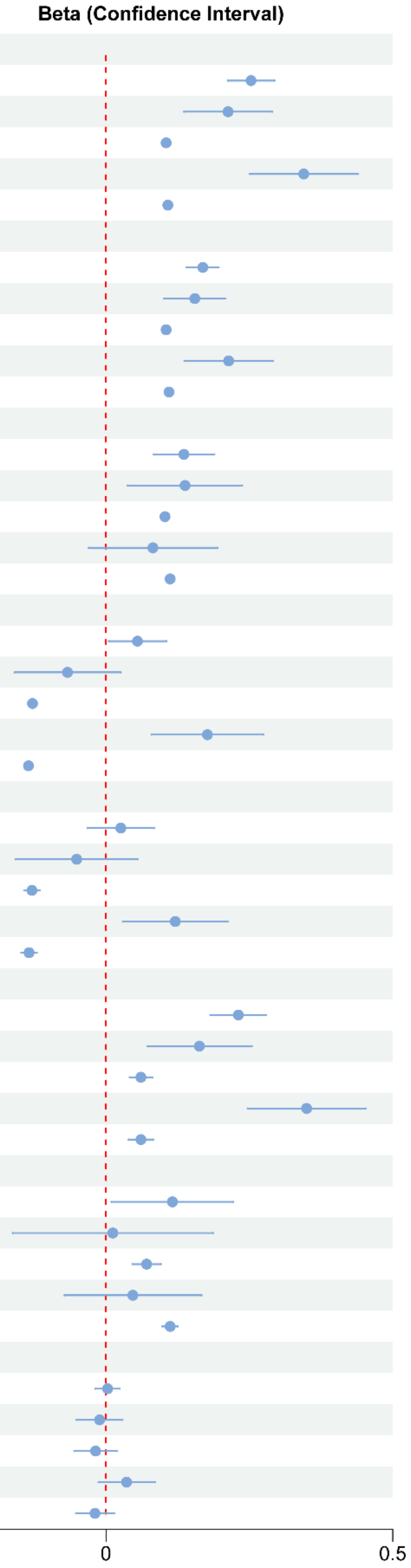

**Figure S1. Effects of omega-3 (secondary) on cardiovascular outcomes (a) and related metabolic traits (b) estimated across five mendelian randomization (MR) methods.** nIVs, number of instrumental variables (IVs) used in the analysis. SE, standard error of the effect estimate. The APSS algorithm employs considerably fewer instrumental variables because it restricts analysis to SNPs included in the HapMap 3 reference panel (See **Methods**).

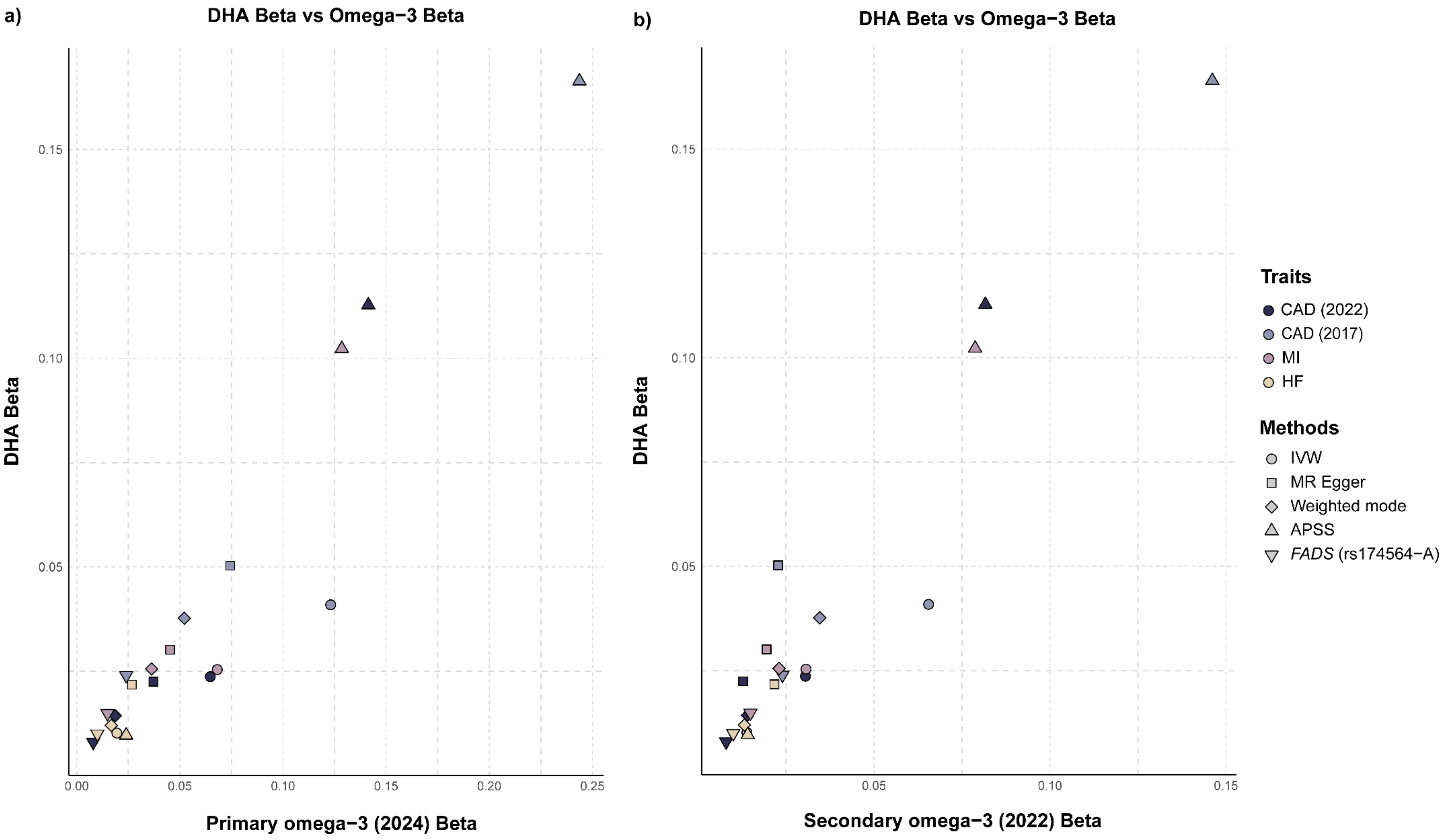

**Figure S2. Comparison of MR results using DHA and omega-3 datasets.** Panels a and b display the MR results using DHA dataset alongside primary omega-3 (Nature 2024) and secondary omega-3 (Plos Biology 2022) datasets, respectively. Each point represents a cardiovascular outcome analyzed with different MR methods. Shape indicates the MR method used, and fill color corresponds to the specific cardiovascular trait. Beta estimates on the x-axis represent effect sizes using omega-3 as the exposure; y-axis values represent the corresponding effect sizes using DHA as the exposure.

a)

| Clinical exposure | Method | nIVs | Beta | SE | *P*-value |
|---|---|---|---|---|---|
| CAD (2022) (181, 522 cases, 984, 168 controls) | | | | | |
| | IVW | 236 | −0.052 | 0.026 | 4.70e−02 |
| | MR Egger | 236 | −0.134 | 0.068 | 4.89e−02 |
| | Weighted mode | 236 | 0.04 | 0.103 | 6.98e−01 |
| | APSS | 206 | −0.019 | 0.035 | 5.88e−01 |
| CAD (2017) (34, 541cases, 261, 984 controls) | | | | | |
| | IVW | 221 | 0.019 | 0.023 | 4.09e−01 |
| | MR Egger | 221 | −0.09 | 0.062 | 1.47e−01 |
| | Weighted mode | 221 | −0.013 | 0.045 | 7.73e−01 |
| | APSS | 176 | 0.059 | 0.031 | 6.04e−02 |
| Myocardial Infarction (61, 505 cases, 577, 716 controls) | | | | | |
| | IVW | 84 | −0.033 | 0.044 | 4.58e−01 |
| | MR Egger | 84 | −0.304 | 0.11 | 7.05e−03 |
| | Weighted mode | 84 | −0.047 | 0.098 | 6.29e−01 |
| | APSS | 76 | −0.009 | 0.049 | 8.46e−01 |
| Heart Failure (47, 309 cases, 930, 014 controls) | | | | | |
| | IVW | 4 | 0.077 | 0.193 | 6.90e−01 |
| | MR Egger | 4 | 0.046 | 1.17 | 9.72e−01 |
| | Weighted mode | 4 | 0.11 | 0.283 | 7.24e−01 |
| | APSS | 5 | −0.151 | 0.244 | 5.38e−01 |
| Atrial Fibrillation (60, 620 cases, 970, 216 controls) | | | | | |
| | IVW | 186 | −0.001 | 0.029 | 9.58e−01 |
| | MR Egger | 186 | 0.073 | 0.074 | 3.25e−01 |
| | Weighted mode | 186 | 0.03 | 0.053 | 5.66e−01 |
| | APSS | 150 | −0.045 | 0.038 | 2.33e−01 |
| Stroke (40, 585 cases, 406, 111 controls) | | | | | |
| | IVW | 8 | −0.218 | 0.227 | 3.37e−01 |
| | MR Egger | 8 | −0.706 | 2.4 | 7.79e−01 |
| | Weighted mode | 8 | −0.134 | 0.236 | 5.87e−01 |
| | APSS | 9 | −0.357 | 0.318 | 2.61e−01 |
| Ischemic Stroke (4, 217 cases, 406, 111 controls) | | | | | |
| | IVW | 9 | −0.288 | 0.238 | 2.27e−01 |
| | MR Egger | 9 | −3.608 | 1.53 | 5.04e−02 |
| | Weighted mode | 9 | −0.058 | 0.171 | 7.42e−01 |
| | APSS | 7 | −0.638 | 0.435 | 1.42e−01 |

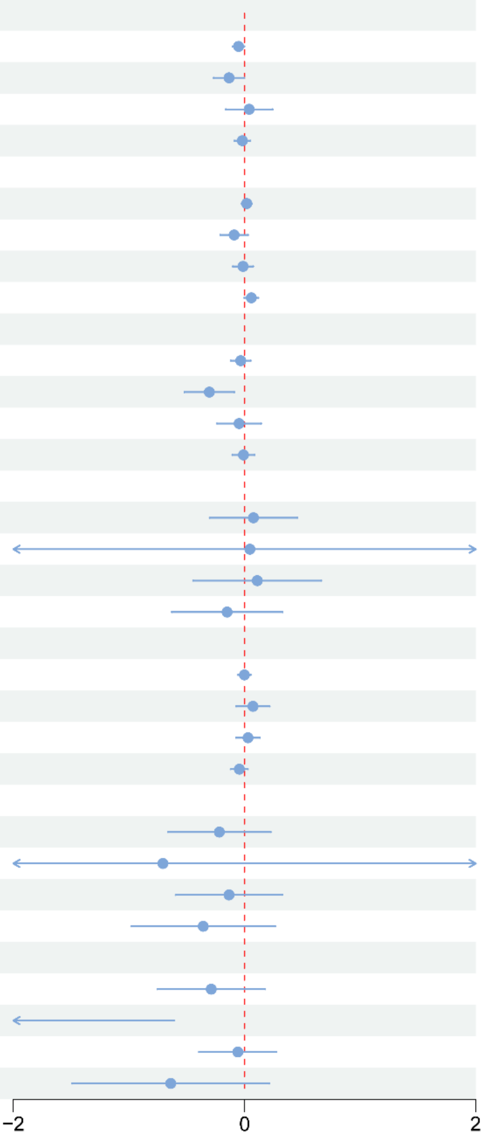

**b)**

| Clinical exposure | Method | nIVs | Beta | SE | *P*-value |
|---|---|---|---|---|---|
| CAD (2022) (181, 522 cases, 984, 168 controls) | | | | | |
| | IVW | 243 | −0.042 | 0.031 | 1.75e−01 |
| | MR Egger | 243 | −0.166 | 0.084 | 4.93e−02 |
| | Weighted mode | 243 | 0.181 | 0.073 | 1.37e−02 |
| | APSS | 208 | −0.007 | 0.038 | 8.55e−01 |
| CAD (2017) (34, 541cases, 261, 984 controls) | | | | | |
| | IVW | 224 | −0.018 | 0.026 | 4.75e−01 |
| | MR Egger | 224 | −0.085 | 0.07 | 2.25e−01 |
| | Weighted mode | 224 | 0.085 | 0.039 | 3.00e−02 |
| | APSS | 180 | 0.034 | 0.039 | 3.74e−01 |
| Myocardial Infarction (61, 505 cases, 577, 716 controls) | | | | | |
| | IVW | 88 | 0.01 | 0.045 | 8.31e−01 |
| | MR Egger | 88 | 0.168 | 0.132 | 2.07e−01 |
| | Weighted mode | 88 | 0.156 | 0.078 | 4.70e−02 |
| | APSS | 73 | −0.014 | 0.066 | 8.29e−01 |
| Heart Failure (47, 309 cases, 930, 014 controls) | | | | | |
| | IVW | 7 | −0.014 | 0.17 | 9.36e−01 |
| | MR Egger | 7 | −0.222 | 0.943 | 8.23e−01 |
| | Weighted mode | 7 | −0.155 | 0.294 | 6.16e−01 |
| | APSS | 8 | 0.134 | 0.277 | 6.27e−01 |
| Atrial Fibrillation (60, 620 cases, 970, 216 controls) | | | | | |
| | IVW | 188 | −0.094 | 0.034 | 5.57e−03 |
| | MR Egger | 188 | 0.033 | 0.087 | 7.10e−01 |
| | Weighted mode | 188 | −0.049 | 0.061 | 4.24e−01 |
| | APSS | 150 | −0.07 | 0.046 | 1.33e−01 |
| Stroke (40, 585 cases, 406, 111 controls) | | | | | |
| | IVW | 9 | −0.143 | 0.174 | 4.14e−01 |
| | MR Egger | 9 | −0.966 | 1.91 | 6.29e−01 |
| | Weighted mode | 9 | −0.257 | 0.305 | 4.24e−01 |
| | APSS | 9 | −0.111 | 0.259 | 6.69e−01 |
| Ischemic Stroke (4, 217 cases, 406, 111 controls) | | | | | |
| | IVW | 11 | −0.191 | 0.155 | 2.18e−01 |
| | MR Egger | 11 | −1.601 | 1.228 | 2.25e−01 |
| | Weighted mode | 11 | −0.323 | 0.29 | 2.91e−01 |
| | APSS | 7 | −0.548 | 0.253 | 3.02e−02 |

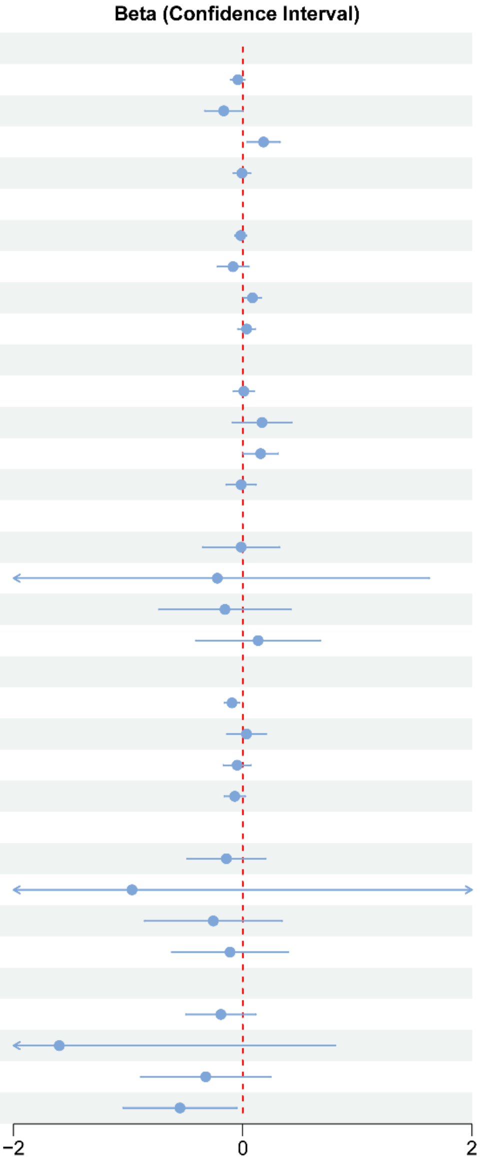

**Figure S3. Reverse MR results using cardiovascular events as the exposure and primary (a) and secondary (b) omega-3 datasets as the outcome, respectively.** nIVs, number of instrumental variables used in the MR analysis. SE, standard error of the effect estimate.

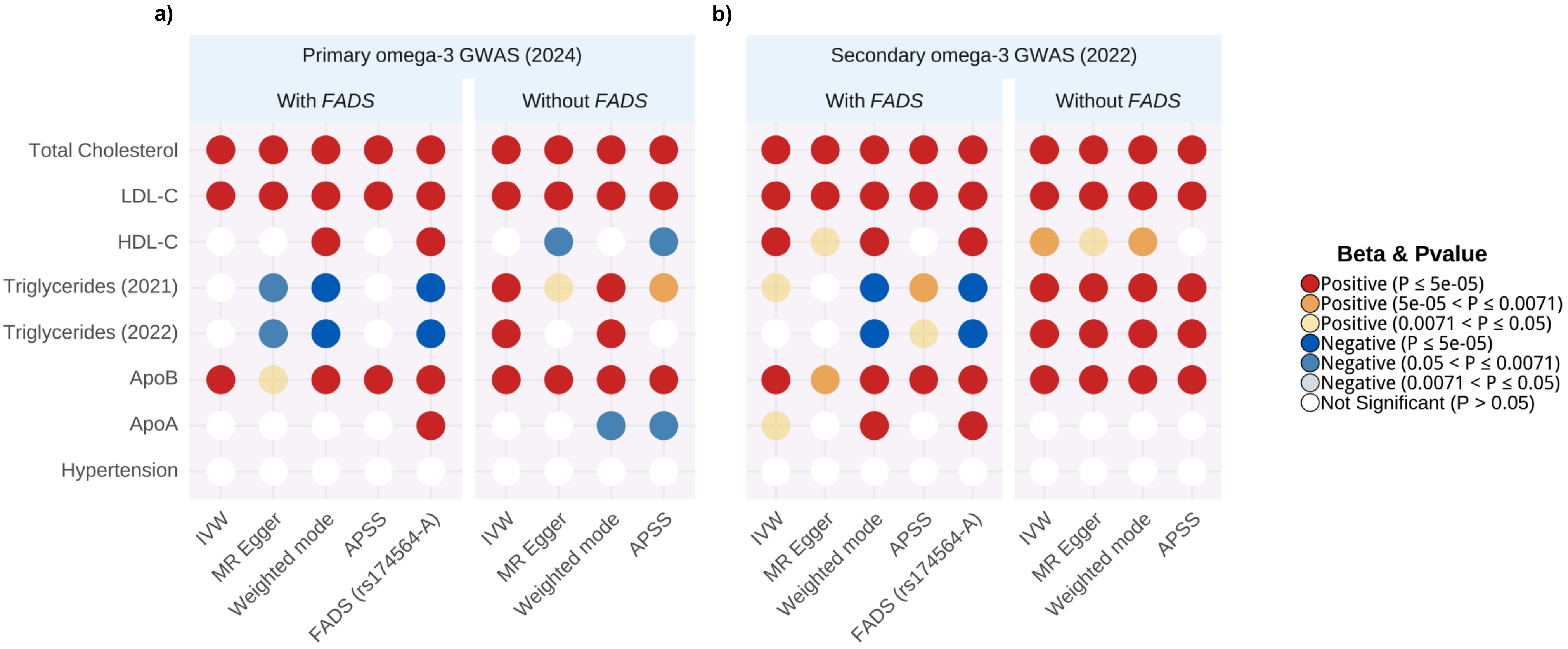

**Figure S4. Effects of omega-3 on metabolic traits with or without inclusion of variants in the *FADS* gene cluster.** (a) Primary omega-3 GWAS derived from a multi-cohort study was used as the exposure. (b) Secondary omega-3 GWAS derived from UK Biobank (2022) was used as the exposure. The *FADS* gene cluster was defined as a region in hg19: chr11:61,067,097 - 62,134,826. The Bonferroni-adjusted significance threshold was set to be 0.0071 (0.05/7).

| Exposure | Outcome | Method | nIVs | Beta | SE | *P* - Value |
|---|---|---|---|---|---|---|
| Primary omega−3 (2024) | LDL-C | IVW | 18 | 0.024 | 0.011 | 2.64E-02 |
| Secondary omega−3 (2022) | LDL-C | IVW | 42 | 0.034 | 0.007 | 3.31E-07 |
| **Combined** | | | | **0.031** | **0.006** | **3.45E-08** |
| Primary omega−3 (2024) | LDL-C | MR Egger | 18 | 0.047 | 0.051 | 3.70E-01 |
| Secondary omega−3 (2022) | LDL-C | MR Egger | 42 | 0.063 | 0.029 | 3.64E-02 |
| **Combined** | | | | **0.059** | **0.025** | **1.90E-02** |
| Primary omega−3 (2024) | LDL-C | Weighted mode | 18 | 0.014 | 0.024 | 5.65E-01 |
| Secondary omega−3 (2022) | LDL-C | Weighted mode | 42 | 0.045 | 0.016 | 7.10E-03 |
| **Combined** | | | | **0.036** | **0.013** | **7.00E-03** |
| Primary omega−3 (2024) | LDL-C | APSS | 16 | 0.028 | 0.017 | 9.98E-02 |
| Secondary omega−3 (2022) | LDL-C | APSS | 26 | 0.028 | 0.010 | 5.68E-03 |
| **Combined** | | | | **0.028** | **0.009** | **1.00E-03** |

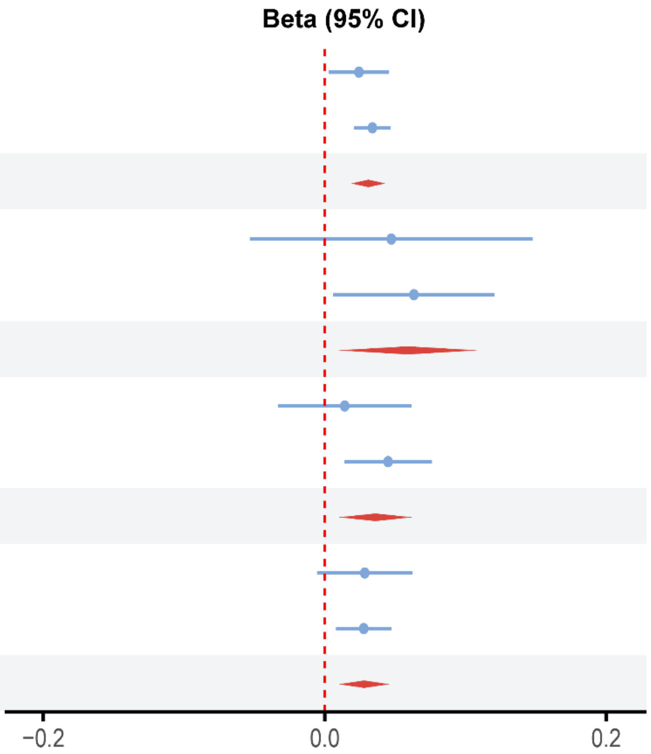

**Figure S5. Causal effect estimates of omega-3 fatty acids on LDL-C levels using different MR methods.** nIVs, number of instrumental variables used in the analysis. SE, standard error of the effect estimate. Here, only genetic variants significantly associated with omega-3 levels (P < 5.00E-08), but not associated with LDL-C levels (P > 0.05), were included as instrumental variables in the MR analyses.

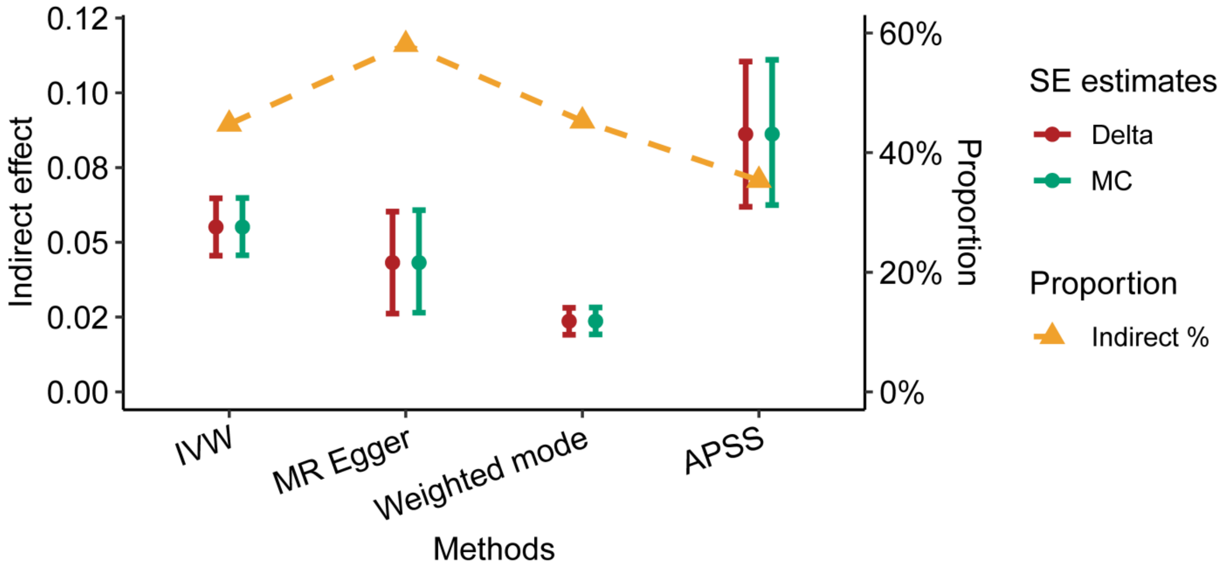

**Figure S6. Mediation analysis of the effect of omega-3 fatty acids on CAD risk through LDL-C.** Indirect effect estimates of omega-3 on CAD through LDL-C using 4 MR methods. Error bars represent 95% CIs calculated using the Delta method (red) and Monte Carlo method (green). Yellow triangles represent proportion of the indirect effect relative to the total effect across 4 MR methods.

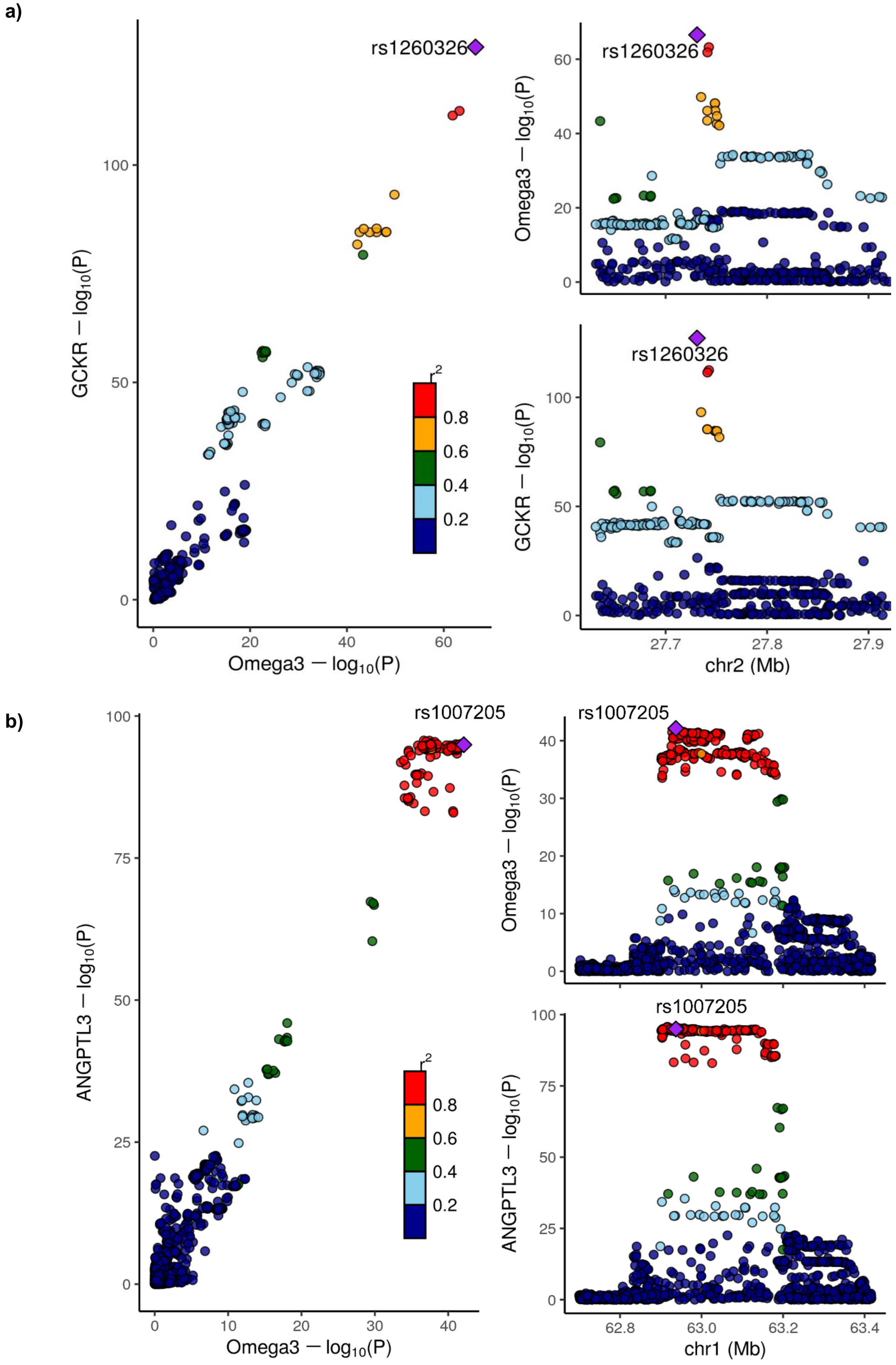

a)
rs1260326
GCKR − log10(P)
Omega3 − log10(P)
r2
0.8
0.6
0.4
0.2
chr2 (Mb)
b)
rs1007205
ANGPTL3 − log10(P)
chr1 (Mb)

c)

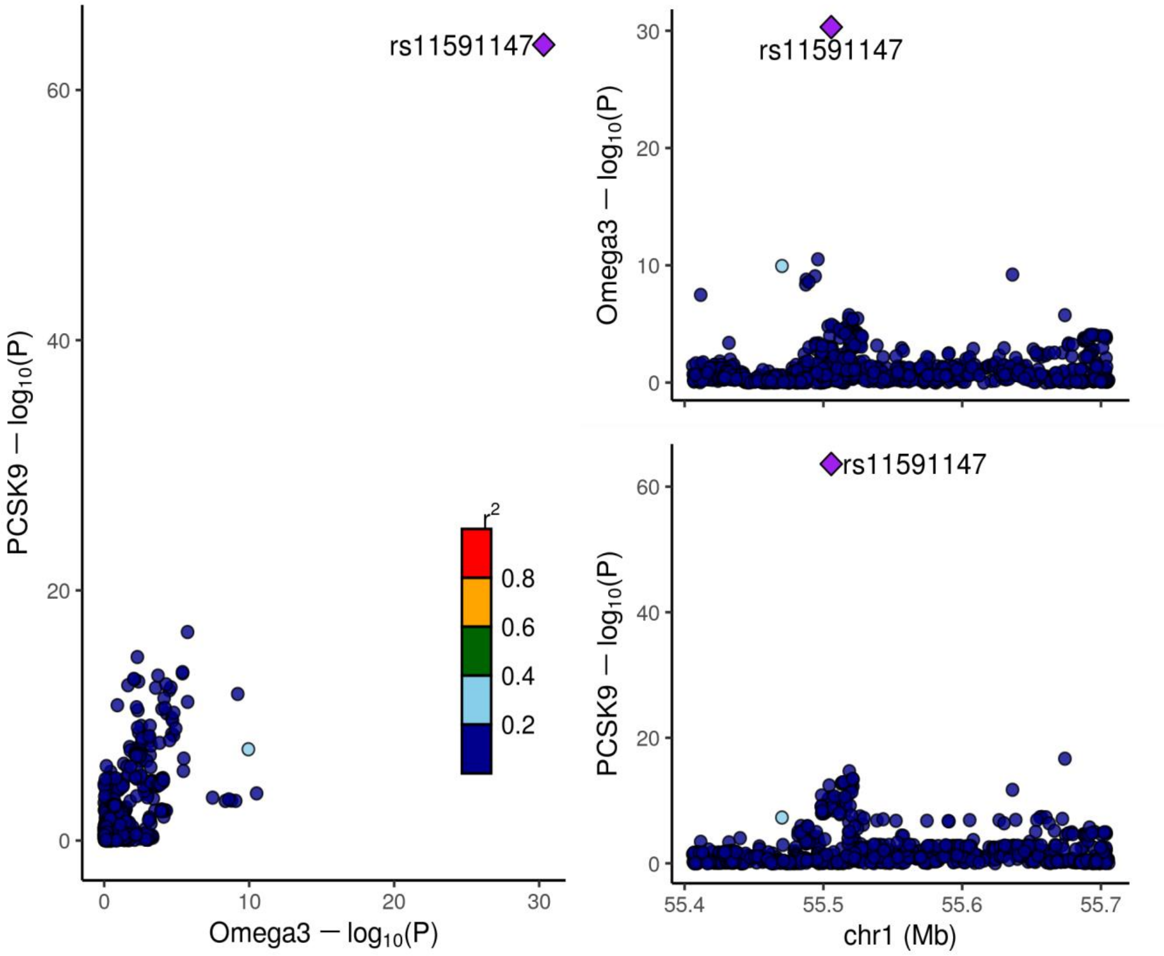
rs11591147
PCSK9 − log10(P)
Omega3 − log10(P)
0
10
20
30
40
60
r2
0.8
0.6
0.4
0.2
chr1 (Mb)
55.4
55.5
55.6
55.7

d)

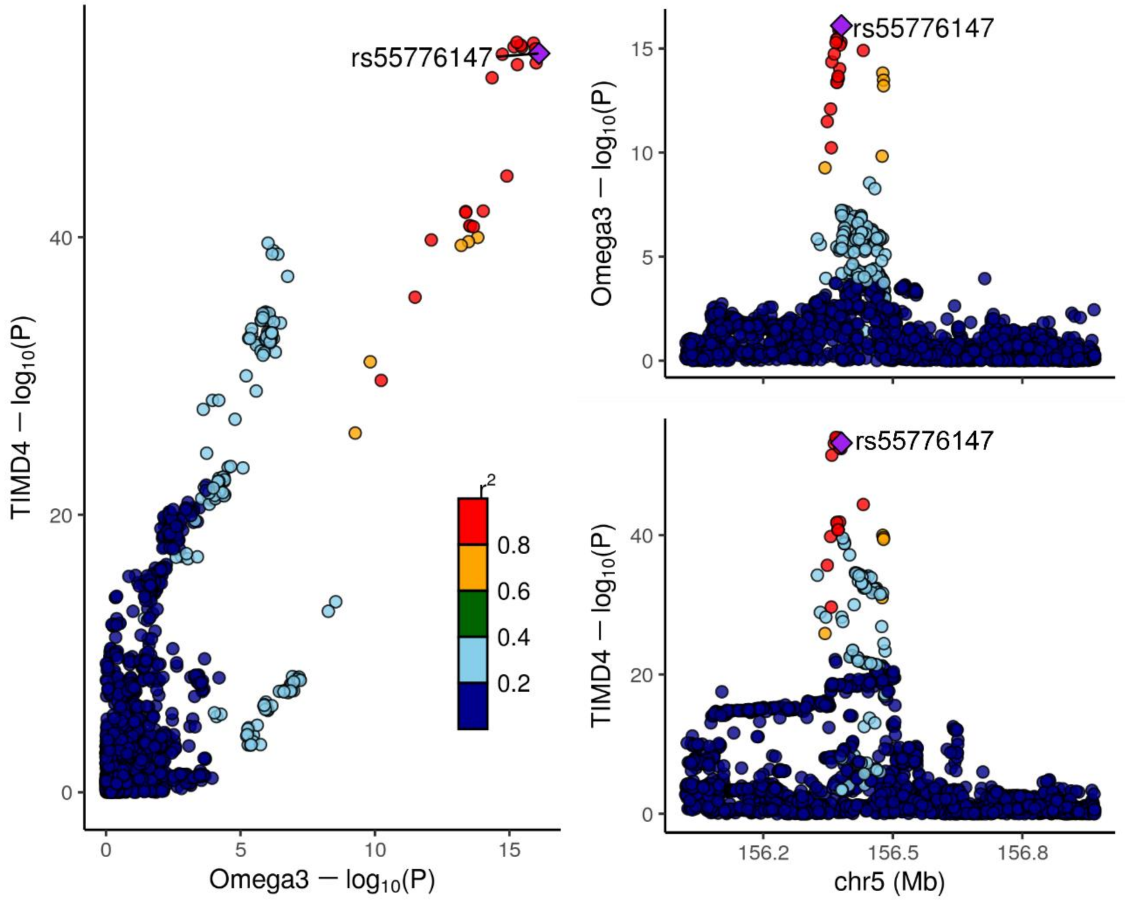
rs55776147
TIMD4 − log10(P)
Omega3 − log10(P)
0
5
10
15
20
40
r2
0.8
0.6
0.4
0.2
chr5 (Mb)
156.2
156.5
156.8

**e)**

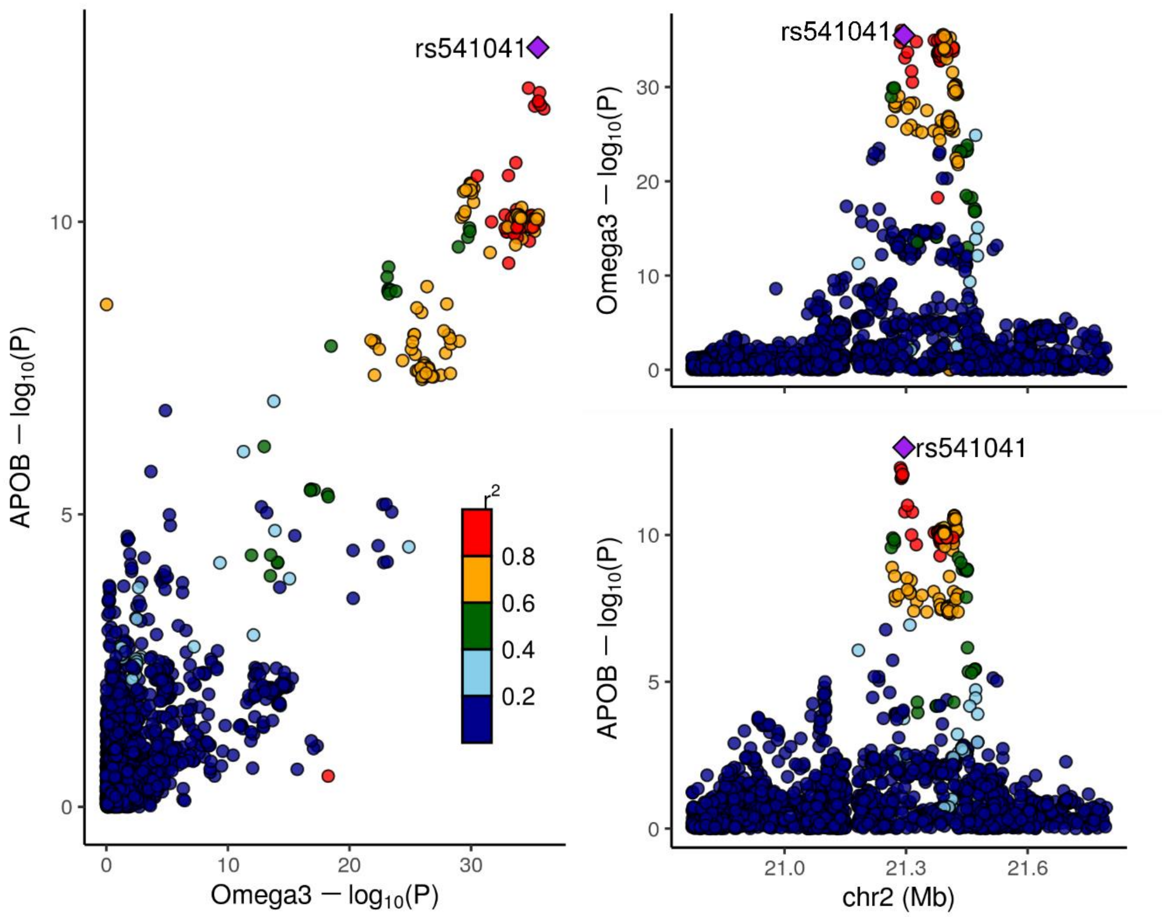

**Figure S7. Identification of plasma proteins associated with omega-3 levels based on the primary omega-3 GWAS.** (a–e) Colocalization between omega3 fatty acid and pQTL association signals. For each locus, the left panel shows an XY scatter plot comparing −log10 P-values for each variant from the GWAS or pQTL datasets; the right panel shows the regional association plot. Five plasma proteins were found significantly associated with the omega-3 levels: (a) GCKR, (b) ANGPTL3, (c) PCSK9, (d) TIMD4, (e) ApoB.

**a)**

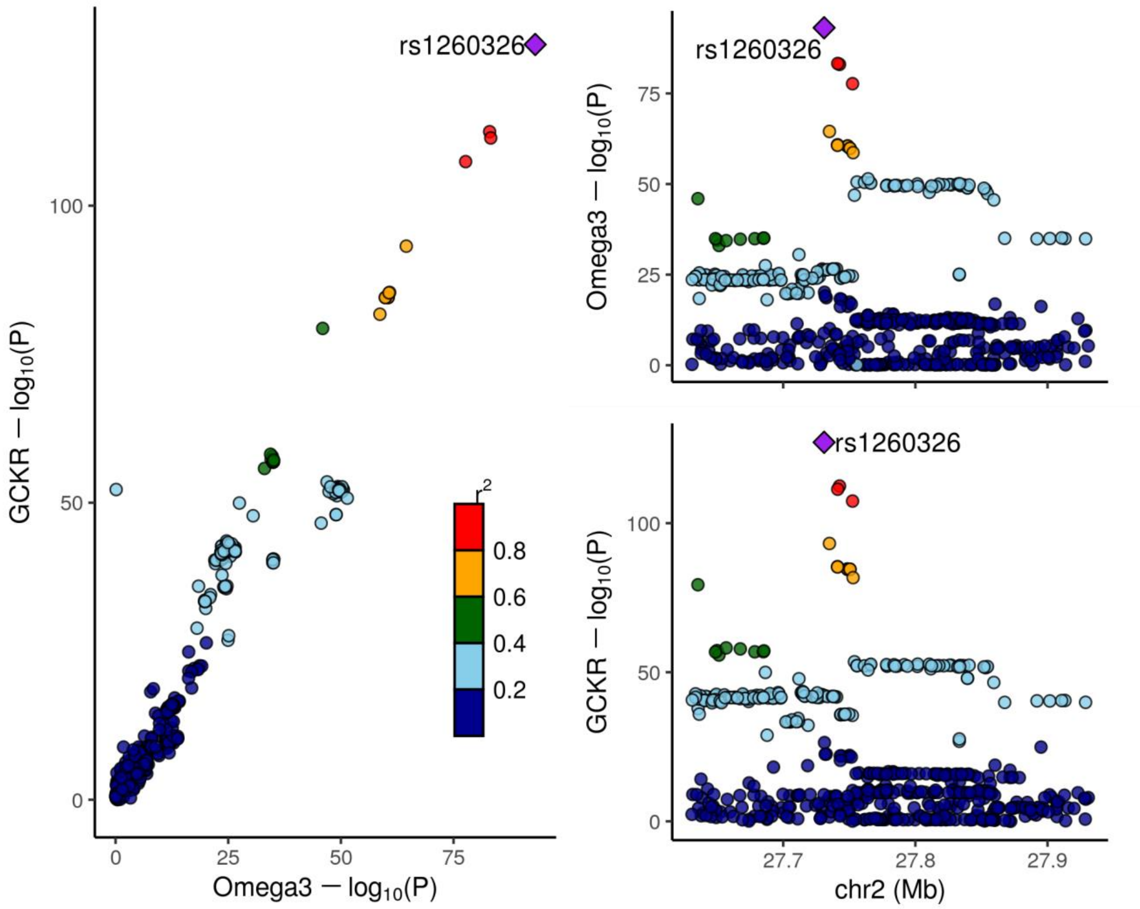

rs1260326
GCKR — $\log_{10}$(P)
Omega3 — $\log_{10}$(P)
$r^2$
0.8
0.6
0.4
0.2
Omega3 — $\log_{10}$(P)
GCKR — $\log_{10}$(P)
27.7
27.8
27.9
chr2 (Mb)

**b)**

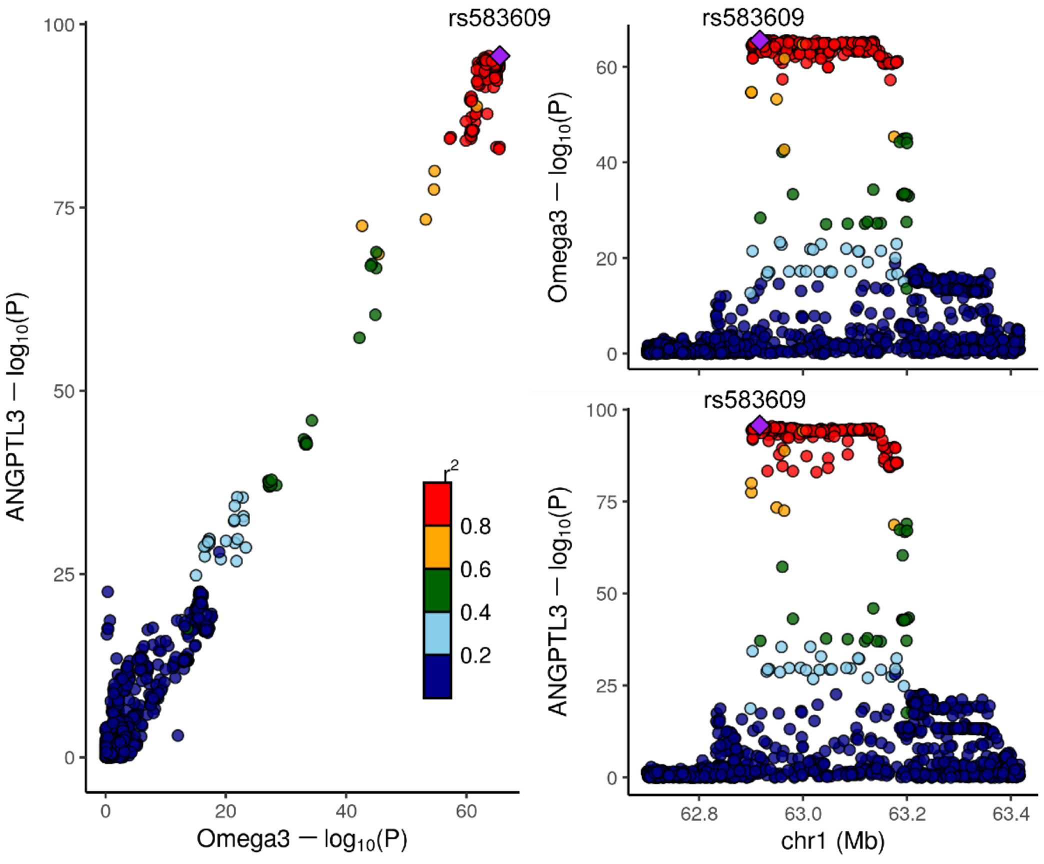

rs583609
ANGPTL3 — $\log_{10}$(P)
Omega3 — $\log_{10}$(P)
$r^2$
0.8
0.6
0.4
0.2
Omega3 — $\log_{10}$(P)
ANGPTL3 — $\log_{10}$(P)
62.8
63.0
63.2
63.4
chr1 (Mb)

**c)**

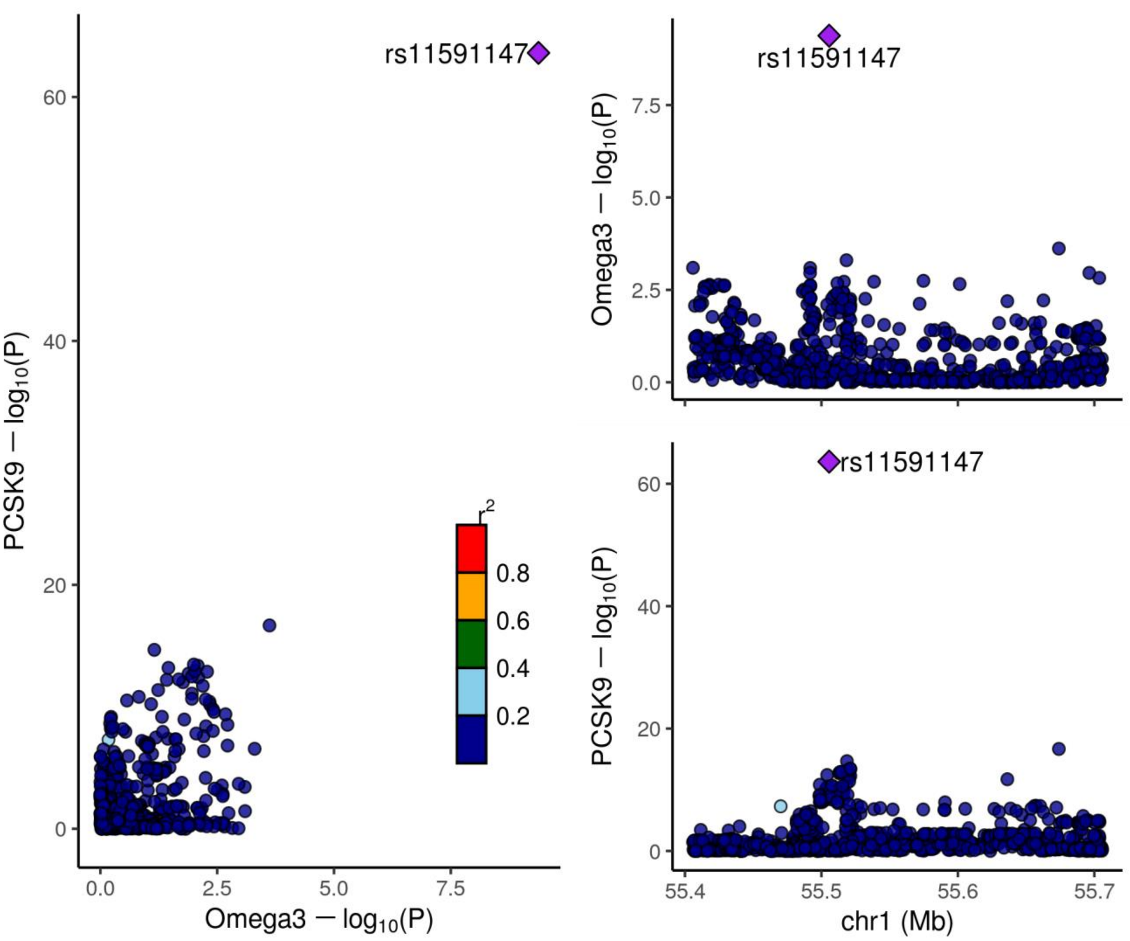
rs11591147
PCSK9 − log10(P)
Omega3 − log10(P)
r2
0.8
0.6
0.4
0.2
chr1 (Mb)
55.4
55.5
55.6
55.7

**d)**

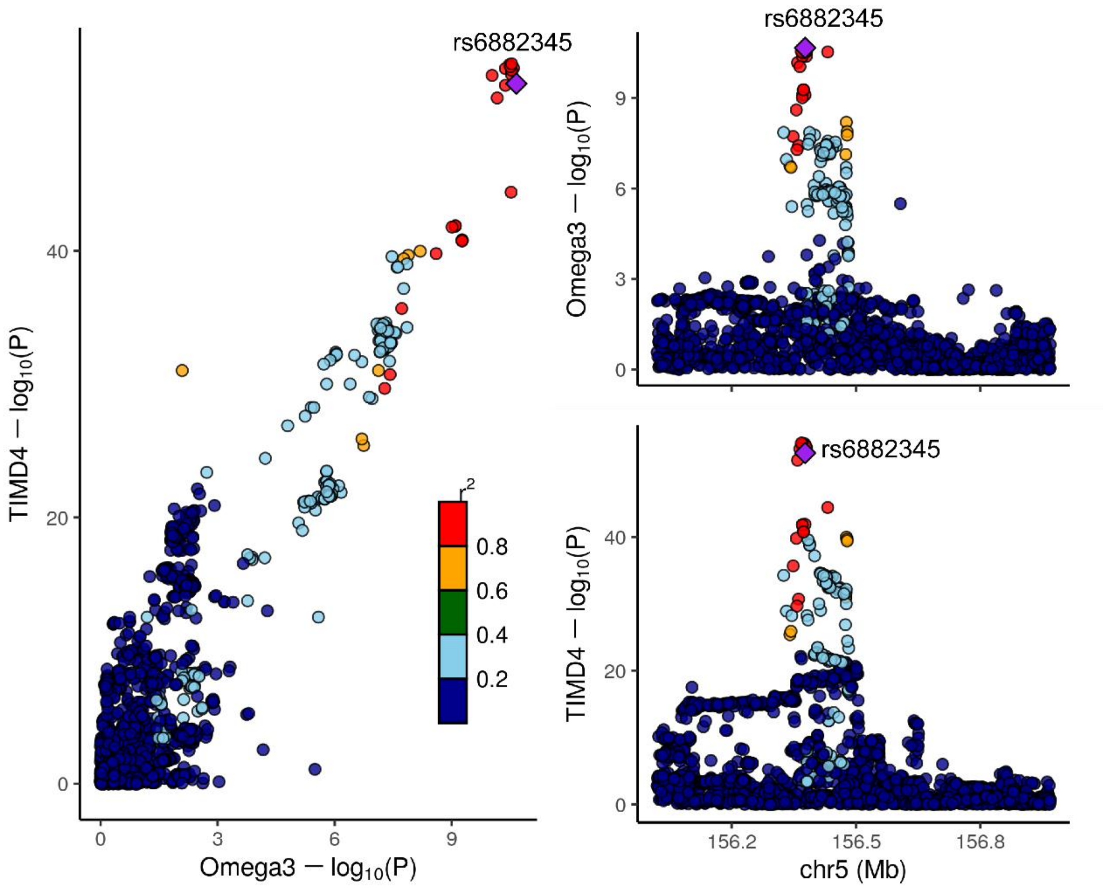
rs6882345
TIMD4 − log10(P)
Omega3 − log10(P)
r2
0.8
0.6
0.4
0.2
chr5 (Mb)
156.2
156.5
156.8

**e)**

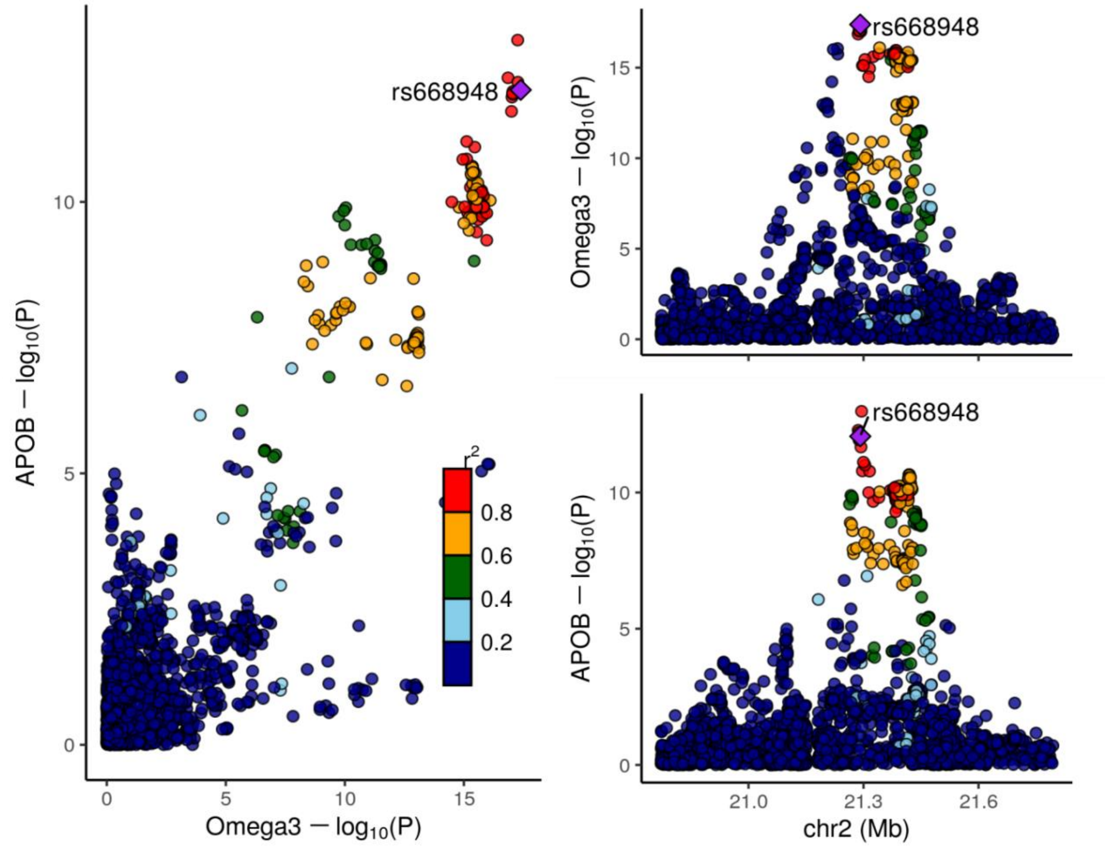

**Figure S8. Identification of plasma proteins associated with omega-3 levels based on the secondary omega-3 GWAS.** (a–e) Colocalization between omega3 fatty acid and pQTL association signals. For each locus, the left panel shows an XY scatter plot comparing −log10 P-values for each variant from the GWAS or pQTL datasets; the right panel shows the regional association plot. Five plasma proteins were found significantly associated with the omega-3 levels: (a) GCKR, (b) ANGPTL3, (c) PCSK9, (d) TIMD4, (e) ApoB
